# Single-cell immune profiling of third trimester pregnancies defines importance of chemokine receptors and prevalence of CMV-induced NK cells in the periphery and decidua

**DOI:** 10.1101/2025.03.24.25324489

**Authors:** Jennifer R Habel, Thi H O Nguyen, Lilith F Allen, Ruth R Hagen, Lukasz Kedzierski, E Kaitlynn Allen, Xiaoxiao Jia, Shihan Li, Ilariya Tarasova, Anastasia A. Minervina, Mikhail V. Pogorelyy, Philippa M Saunders, Allison Clatch, Maximilien Evrard, Calvin Xu, Hui-Fern Koay, Md Abdullah-Al-Kamran Khan, Natasha de Alwis, Laura K Mackay, Alexander D Barrow, Celia Douros, Theo Karapanagiotidis, Suellen Nicholson, Katherine Bond, Deborah A Williamson, Martha Lappas, Susan Walker, Natalie J Hannan, Andrew G Brooks, Jan Schroeder, Jeremy Chase Crawford, Paul G Thomas, Louise C Rowntree, Katherine Kedzierska

## Abstract

Human pregnancy presents a unique physiological state that allows for growth of an antigenically dissimilar foetus and requires specific adaptations of the immune system. The immune system plays an important role in establishing and maintaining successful pregnancy, yet deep understanding of immunological responses in pregnancy is lacking. To provide in-depth understanding of the immunological landscape of pregnancy, with a focus on NK cells, we used high-throughput cell surface proteome screening of >350 markers and scRNAseq with 130 CITE-seq antibodies to identify key differentially expressed molecules and investigated their potential roles in altered immunity. We identified skewing in NK cell subsets towards higher frequencies of CD56^bright^ cells caused by a reduced number of CD56^dim^ cells in the peripheral blood during pregnancy and provide evidence for a role of chemokine receptor, CX3CR1, in NK cell activation. In addition, we defined a new cytomegalovirus (CMV)-induced decidual NK cell population in CMV^+^ pregnancies with tissue-residency markers. Overall, our data provide fundamental knowledge into how NK cell immunity is altered in pregnancy, and key knowledge needed to inform vaccine and therapeutic strategies to manage infections or pregnancy complications.

## INTRODUCTION

Human pregnancy presents a unique physiological state that allows for growth of an antigenically dissimilar foetus. While there is partial physical separation of the foetus by amniotic and chorionic membranes, successful human pregnancy requires a tightly regulated immune system to remain non-reactive toward foetal antigens. To achieve this, there is a characteristic bias towards type II or tolerogenic immunity during pregnancy (reviewed in^1^).

Despite the tolerogenic state of immunity during gestation, specific activation is still required to establish the pregnancy and during labour and parturition. Natural killer (NK) cells have a well-established role in placentation and establishment of foetal vascular supply, which rely on maternal NK cell activating killer cell immunoglobulin-like receptors (KIRs) interacting with HLA-C on foetal-derived trophoblast cells^2^. Impairments in this interaction can result in life-threatening hypertensive disorder pre-eclampsia, often leading to major organ and cardiovascular injury^3^. Furthermore, uterine NK cells secrete IFN-γ in response to invading trophoblast, causing widening of spiral arteries which is essential for optimal blood flow and nutrient exchange in healthy pregnancies^4,5^.

In addition to NK cell involvement at the maternal-foetal interface, maternal peripheral NK cells undergo phenotypic changes, which are, at least in part, caused by signalling from trophoblasts^6^. Increase in HLA-DR^+^ NK cells during healthy pregnancy has been described, and more specifically, increased expression of activation markers CD38, NKp46, PD-1 and CD27 within the CD56^dim^ NK cell subset can be found^7,8^. Additionally, in first trimester pregnancies (6-12 weeks), there is an increased frequency of IL-10^+^ CD56^bright^ NK cells in the blood compared to non-pregnant women^9^. Furthermore, the importance of balanced NK cell activation during pregnancy is exemplified in recurrent pregnancy loss, where women with recurrent pregnancy loss have increased NK cell cytolytic activity compared to women with no pregnancy loss, suggesting that NK cell cytotoxic functions need to be well regulated for healthy and full-term pregnancy^10^.

Despite clear evidence that NK cells play an important role in pregnancy, fundamental knowledge is lacking surrounding the mechanisms underpinning their altered phenotypes and functions, especially within peripheral blood. This is particularly relevant given that pregnancy is often an exclusion criterion for clinical trials whereby several medications and vaccines have unknown safety and efficacy in pregnancy. To provide in-depth understanding of the immunological landscape of pregnancy, with a focus on NK cells, we used high-dimensional approaches to identify key differentially expressed molecules and investigated their potential roles in altered immune functionality. We identified a skewing of NK cell subsets towards higher frequencies of CD56^bright^ cells caused by a reduced number of CD56^dim^ cells in the peripheral blood during pregnancy and provide evidence for a role of chemokine receptor, CX3CR1, in NK cell activation. In addition, we defined a new cytomegalovirus (CMV)-induced decidual NK cell population in CMV^+^ pregnancies with tissue-residency markers. Overall, our data provide fundamental knowledge into how NK cell immunity is altered in pregnancy, and key knowledge needed to inform vaccine and therapeutic strategies to manage infections or pregnancy complications.

## RESULTS

### Pregnancy alters surface proteome of lymphoid and myeloid immune subsets

The immune system plays an important role in establishing and maintaining successful pregnancy, yet deep understanding of immunological responses in pregnancy is lacking. Understanding pregnancy-specific immunology is key for the rational design of vaccines, immunotherapies and biomarkers for safe pregnancy outcomes. To define in-depth the surface proteome of lymphoid and myeloid immune subsets during pregnancy, we screened 360 cell surface proteins using a Massively Parallel Cytometric (MPC) approach (Fig. 1A). We labelled pregnancy-matched maternal PBMCs and decidua samples, pregnancy-matched umbilical cord blood and non-pregnant PBMCs with unique fluorescent barcodes and a backbone of lineage and activation markers (Supplementary Tables 1-2). Samples were then multiplexed and labelled with individual PE-conjugated mAbs for parallel screening. Individual samples were deconvoluted based on fluorescent barcodes and manually gated to identify NK cells, CD8^+^ T cells, CD4^+^ T cells, γδ T cells, B cells and monocytes (Fig. 1B, Supplementary Fig. 1). Across the six immune cell subsets, 110 unique proteins were differentially expressed between pregnant and non-pregnant groups (Fig 1C; Supplementary Fig. 2).

**Figure 1.**
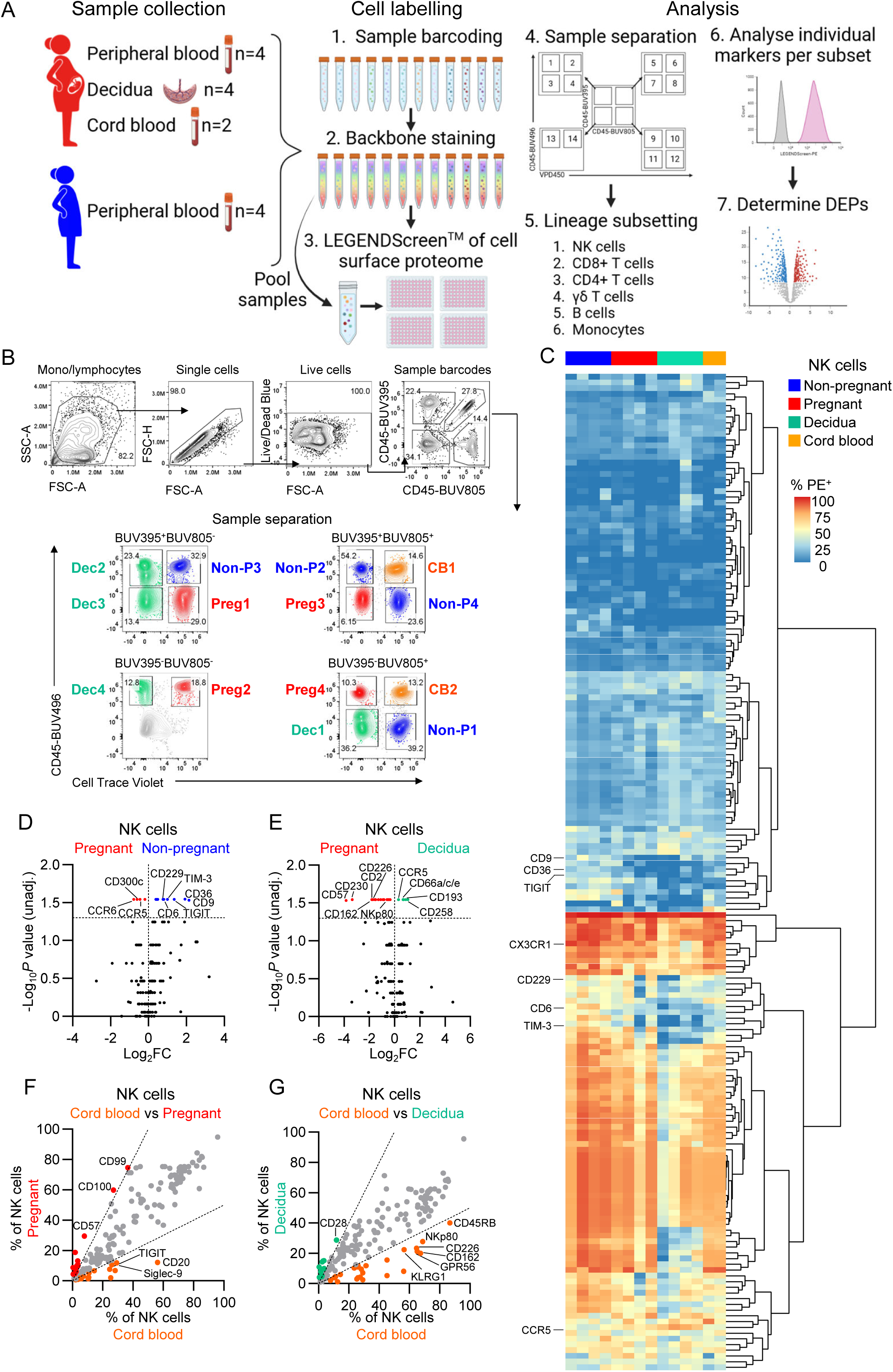
Surface proteome profiling of immune cells from pregnant and non-pregnant women peripheral blood, cord blood and decidua. (A) Schematic representation of the massively parallel cytometry experiment workflow. (B) Representative FACS plots of the gating strategy to deconvolute individual samples based on unique fluorescent barcoding. (C) Heatmap with hierarchical clustering of columns showing frequencies of marker-PE expression on NK cells from pregnant and non-pregnant peripheral blood, and decidua. Markers expressed on >10% of cells in at least one sample were included for analysis (174 of 342 markers). (D,E) Volcano plots depicting differential protein expression in non-pregnant vs pregnant blood NK cells (D) and decidual vs pregnant blood NK cells (E). Volcano plot horizontal dashed line indicates log_10_ p-value cutoff of 1.3 (p<0.05). (F,G) Scatter plot of average percent of cells expressing for cord blood vs pregnant peripheral (F) and cord blood vs decidual (G) NK cells. Scatter plot dashed lines indicate a log_2_FC cutoff of 1.

NK cells play important roles in pregnancy and have been implicated in the essential process of placentation^2,3^, while dysregulation of NK cells is linked to pregnancy complications such as pre-eclampsia^10^. We thus first focused on examining NK cells and identified increased frequencies of migration markers (CCR5, CCR6) and activating receptor CD300c on NK cells but reduced frequencies of NK cells-expressing markers of exhaustion/activation (TIGIT, TIM-3, CD119/IFNGR1, CD337/NKp30, CD229/SLAMF3), scavenger receptors (CD6, CD36), tetraspanin CD9, and adhesion molecule CD31 in pregnant blood compared to non-pregnant blood, but (Fig. 1D). Of these, CD9, CD36, TIGIT and TIM-3 frequencies were reduced by more than 2-fold (log_2_ fold-change (log_2_FC) >1) (Fig. 1D; Supplementary Table 3). CD9 was consistently decreased across CD8^+^, CD4^+^ and γδ T cells, while CD119/IFNGR1 was decreased on γδ T cells and B cells from the pregnant blood (Supplementary Fig. 3A; Supplementary Table 3). Of the markers increased on pregnant NK cells, CD195/CCR5 was also increased on CD8^+^ T cells (Supplementary Fig. 3A). Within the T cell compartment, there were 17, 3 and 13 differentially expressed proteins across CD8^+^, CD4^+^ and γδ T cells, respectively. Notably, migration/adhesion markers CD195/CCR5, CX3CR1, and CD11c/ITGAX were increased for pregnant CD8^+^ T cells, while co-stimulatory molecules CD26 and CD28 were decreased (Supplementary Fig. 3A). In contrast to NK cells and CD8^+^ T cells, there was lower CD195/CCR5 expression on pregnant γδ T cells, with γδ T cells having additional decreases in CD218a/IL18Ra, CD278/ICOS, CD94, CD226/DNAM-1, and CD183/CXCR3 expression (Supplementary Fig. 3A; Supplementary Table 3).

We next investigated differences between pregnant blood and decidua from pregnancy-matched samples. Twenty-three differentially expressed proteins were observed in NK cells between the pregnant blood and decidua, with adhesion molecules (CD99, CD230/prion), activation markers (NKp80, CD226/DNAM-1, CD2), scavenger receptor CD6, anti-migration/inhibitory marker GPR56, migration marker CD162/P-selectin glycoprotein ligand-1 (PSGL-1), and adaptive/CMV-associated NK marker CD57 being higher in the blood, while only CD258/LIGHT was more than 2-fold higher (log_2_FC>1) in decidual NK (dNK) cells (Fig. 1E, Supplementary Table 4). Frequencies of NK cells expressing chemokine receptors CD193/CCR3 and CD195/CCR5 were also higher on dNK but with a lower fold change (Supplementary Table 4). All three proteins (CD193/CCR3, CD195/CCR5, and CD258/LIGHT) were also increased on decidual CD8^+^ and CD4^+^ T cells, while CD195/CCR5 was shared with B cells. CD162/ PSGL-1, CD99, and CD230/prion were higher across all lymphocyte subsets from the pregnant blood, indicating their importance for circulating lymphocytes, but not monocytes (Supplementary Fig. 3B; Supplementary Table 4). Furthermore, CMV-associated marker CD57 was also increased on peripheral γδ T cells, suggesting there may be less adaptive-like NK and γδ T cells within the decidua (Supplementary Fig. 3B; Supplementary Table 4). Markers shared between CD4^+^ and CD8^+^ T cells in the decidua included migration/adhesion markers CD193/CCR3, CD195/CCR5, CD11c, and CXCR7, and Fc-γ receptors (FcRs) CD32 and CD64 (Supplementary Fig. 3B; Supplementary Table 4).

We also analysed differentially expressed markers on NK cells in the relatively immunologically-naïve cord blood. In comparison to pregnant peripheral blood, NK cells in the cord blood had increased CD9 and inhibitory/exhaustion markers siglec-9 and TIGIT (Fig. 1F; Supplementary Table 5). CD9 and siglec-9 were also increased on cord blood monocytes, and CD9 on cord blood B cells (Supplementary Fig. 3C; Supplementary Table 5). Lower CD57 expression was observed on cord blood NK cells and γδ T cells compared to pregnant blood, an indication of a lack of adaptive-like cells in the immunologically naïve neonate (Supplementary Fig. 3C; Supplementary Table 5). When compared to dNK cells, 21 markers were increased on cord blood NK cells, including CD360/IL-21R, CD24, CD230, and CD162/PSGL-1, which were also increased on cord blood CD8^+^, CD4^+^ and γδ T cells (Fig. 1G; Supplementary Fig. 3D; Supplementary Table 6). Furthermore, inhibitory molecule CD200 was increased in all cord blood T cell subsets when compared to pregnant blood or decidual T cells, and adhesion molecule CD49a was increased on cord blood CD4^+^ and CD8^+^ T cells as compared to pregnant blood (Supplementary Fig. 3C,D; Supplementary Table 5,6). Markers increased on cord blood B cells compared to both pregnant blood and decidua included differentiation marker CD10, chemokine receptors CD192/CCR2 and CD123/CCR3, B-1 cell associated markers CD5 and CD148 (Supplementary Fig. 3C,D; Supplementary Table 5,6). There were 66 proteins increased on cord blood monocytes compared to pregnant blood and 34 when compared to the decidua. The top 5 differentially expressed markers increased on cord blood monocytes were CD2, CD15, CD41, CD200R, and CD354 in comparison to pregnant blood, of which CD2, CD15, and CD41 were also within the top 5 when compared to decidual monocytes (Supplementary Fig. 3C,D; Supplementary Table 5,6).

As NK cells play a key role in pregnancy, we further investigated differentially expressed markers TIGIT, CCR5, TIM-3, CD9 and CD36 on NK cells within one flow cytometry panel (Fig. 2A). We performed directly *ex vivo* whole blood staining to validate differential expression of these markers in a larger cohort of pregnant and non-pregnant individuals (Fig. 2B; Supplementary Table 7). We confirmed lower frequencies of CD9^+^ and CD36^+^ NK cells in pregnancy, while there were significantly higher frequencies of CCR5^+^ NK cells (Fig. 2C). Interestingly, dual-staining of CD9 and CD36 revealed that these two markers are highly co-expressed (Fig. 2B,C). TIGIT and TIM-3, however, was not statistically different across the larger cohort (Fig. 2C). Taken together, the whole blood staining of the key differentially expressed surface molecules depicted by the LEGENDScreen showed lower frequences of CD9^+^CD36^+^ NK cells and higher frequencies of CCR5^+^ NK cells during pregnancy, suggesting altered migration patterns and metabolic processes of NK cells in pregnancy.

**Figure 2.**
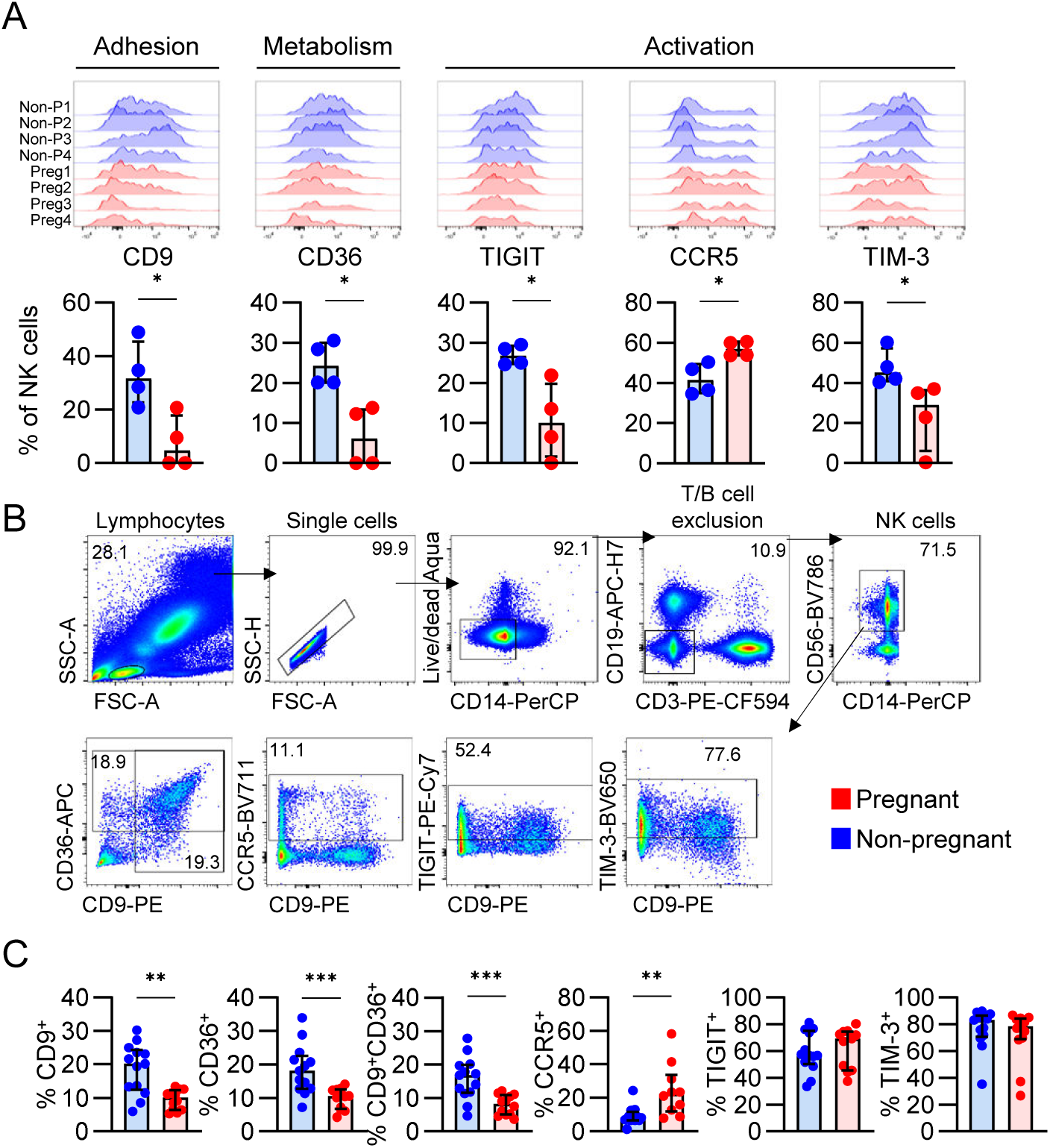
Surface proteome analysis of NK cells from pregnant and non-pregnant blood and decidua. (A) Histograms of selected DE proteins between pregnant and non-pregnant NK cells, and respective scatter plots of the frequency of NK cells expressing each marker. (B) Representative FACS plots showing gating of CD9, CD36, CCR5, TIGIT and TIM-3 on total NK cells and (C) their frequencies from whole blood staining.

### NK cell heterogeneity in peripheral blood and decidua

NK cell features in healthy non-pregnant peripheral blood and across tissues are well-defined. From a pregnancy perspective, studies investigated decidual or uterine NK cells^11–16^. There are, however, limited data exploring NK cells from the peripheral blood of pregnant women at the single-cell resolution. Therefore, we used single-cell RNA sequencing (scRNAseq) with cellular indexing of transcriptomes and epitopes (CITE-seq) to assess RNA expression as well as 130 surface proteins via antibody-derived tags (ADTs) to analyse CD56^+^CD3^-^ NK cells sorted from full-term pregnant PBMCs, pregnancy-matched decidua, and non-pregnant PBMCs (Fig. 3A; Supplementary Figs. 4-5). RNA and protein expression data were used to generate a weighted nearest neighbour uniform manifold approximation projection (WNN-UMAP) of NK cells (Fig. 3B,C; Supplementary Fig. 4B). Following quality control, six unsupervised NK cell clusters were generated from peripheral blood (clusters 0, 4, 6, 7, 9, 12) and seven clusters of dNK cells (clusters 1, 2, 3, 5, 8, 10, 11) (Fig. 3C; Supplementary Fig. 4C-E).

**Figure 3.**
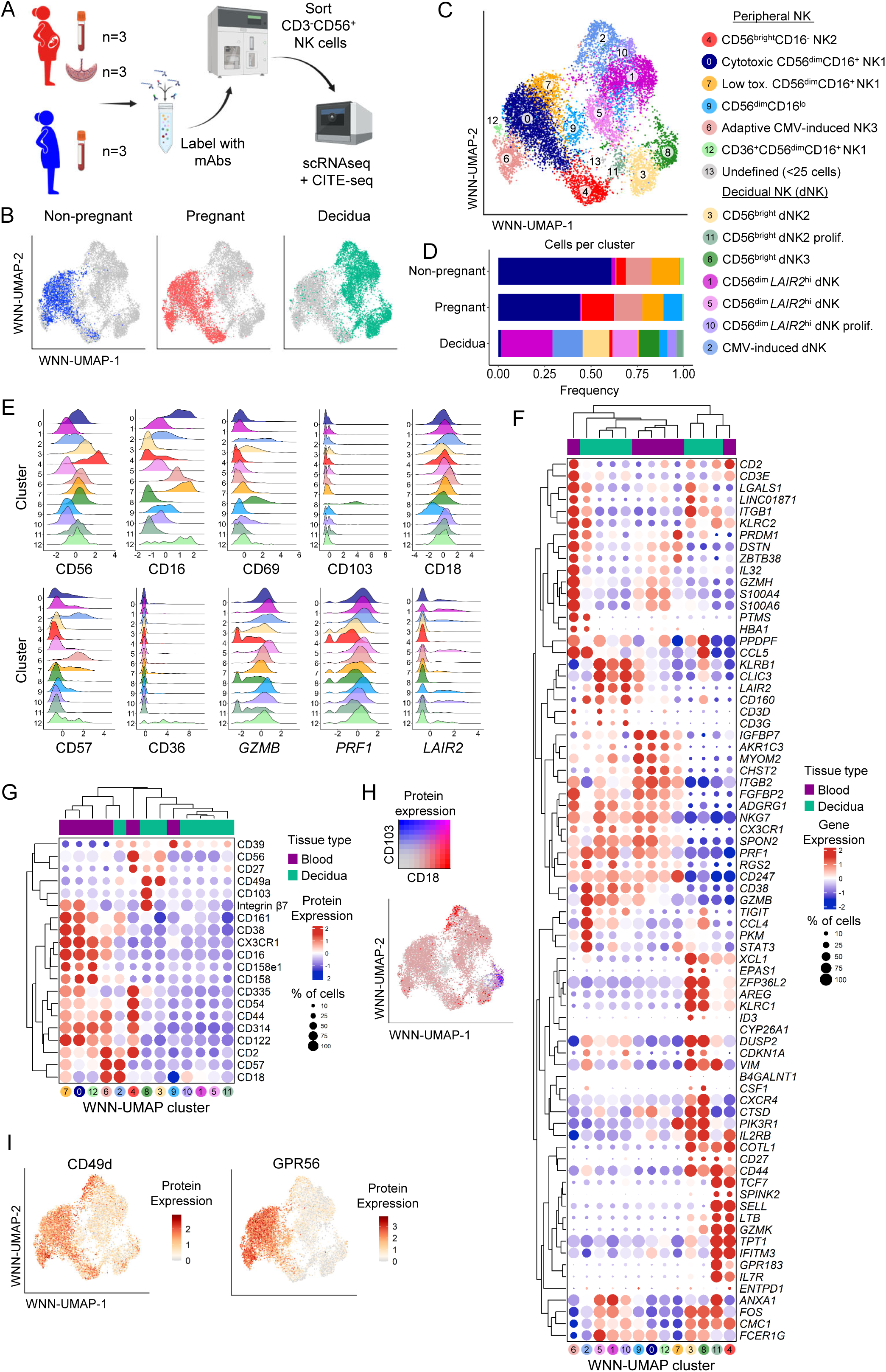
Single cell resolution of NK cell transcriptomes and proteomes in pregnancy. (A) Schematic overview of the experimental approach for scRNAseq on NK cells from pregnant and non-pregnant peripheral blood, and the decidua. (B) WNN-UMAP with NK cells coloured by pregnant, non-pregnant or decidua groups. (C) Weighted nearest neighbour-(WNN-) UMAP of NK cells based on protein and gene expression. (D) The proportion of cells in each WNN-UMAP cluster per group. (E) Histograms depicting the expression levels of selected proteins and genes used to define the WNN-UMAP clusters. (F,G) Hierarchically clustered dot plot of 79 selected genes (F) and 20 selected proteins (G) contributing to distinct NK cell phenotypes observed in each WNN-UMAP cluster. (H) WNN-UMAP of CD18 and CD103 protein co-expression. (I) WNN-UMAP of CD49d and GPR56 CITE-seq expression level.

Clusters were manually annotated using protein expression of canonical NK cell markers CD56 and CD16 to define CD56^bright^CD16^-^ (clusters 3, 4, 8, 11) and CD56^dim^CD16^+^ (clusters 0, 1, 2, 4, 5, 6, 7, 9, 10, 12) (Fig. 3E). We next used 20 surface proteins and 75 genes previously associated with peripheral blood or dNK subsets to further define and validate the NK cell clusters^13,14,17^ (Fig. 3F,G; Supplementary Fig. 4F; Supplementary Table 8). NK cells from peripheral blood were assigned as cytotoxic CD56^dim^/NK1 (clusters 0, 7, 12), CD56^bright^/NK2 (cluster 4), or adaptive CMV-induced/NK3 (cluster 6) (Fig. 3B,C). Of note, cluster 9 did not fall into the peripheral NK1-3 subsets, was CD56^dim^CD16^lo^, and only contained NK cells from one pregnant participant’s blood and decidua (Fig. 3B,C; Supplementary Fig. 4E).

Decidual NK cell clusters were assigned using markers described by Vento-Tormo *et al.* where possible^14^ (Fig. 3B; Supplementary Table 8). Importantly, dNK1-3 subsets were originally described in first trimester decidua and are poorly defined for full-term pregnancies. dNK1 cells are generally distinguished by expression of CD39/*ENTPD1* and killer-cell immunoglobulin-like receptors (KIRs) and absence of CD18/*ITGB2*, while dNK2 and dNK3 cells are CD39^-^CD18^+^, with dNK3 cells also expressing CD103^14^. Interestingly, in our dataset, CD18/*ITGB2* expression was observed across all dNK clusters, although with higher expression detected in clusters 2, 3 and 8, and low CD39/*ENTPD1* expression, indicating the dNK1 subset was not present or underwent a significant phenotypic shift in decidua from full-term pregnancies (Fig. 3F,G). This aligns with Whettlock *et al*., who showed dNK1 cells peaked in the first trimester and diminished over the course of pregnancy^15^. Meanwhile, dNK2 cells (CD18/*ITGB2^+^*) were observed in cluster 3 and dNK3 cells (CD18^+^CD103^+^) in cluster 8 (Fig. 3E,G,H).

Other dNK cells in clusters 1, 5, and 10 were distinguished by their *LAIR2* expression and could not easily be defined by the dNK1-3 subsets (Fig. 3E-G). These clusters were CD56^dim^CD16^+^, but unlike CD56^dim^ cells observed in the peripheral blood, they lacked periphery-associated markers GPR56 and CD49d^18^ (Fig. 3I). Additionally, similar to dNK3 cells, clusters 1, 5 and 10 expressed *KLRB1* and *CLIC3*, however they also expressed genes associated with the peripheral NK1 subset, including *NKG7, CX3CR1, SPON2,* and *FGFBP2* (Fig. 3F). Similarly, cluster 11 had some characteristics of both dNK2 (e.g. *CD2, KLRC2, ANXA1, CD27)* and peripheral NK2 (e.g. *GZMK, XCL1, TCF7, IL7R, IFITM3*) subsets (Fig. 3F). It is possible that these decidua-derived NK cells were circulating through the tissue or recently migrated, thus had characteristics of both circulating and decidual NK cells.

### A fourth dNK subset in CMV-seropositive pregnancies

Both peripheral blood NK and dNK cells formed clusters of CMV-induced adaptive NK cells (clusters 6 and 2, respectively) defined by expression of *KLRC2*, *B3GAT1*/CD57, and *LAG3*/CD223 (LAG-3)^13,17,19^, predominantly observed in donors with a CMV^+^ serostatus (Fig. 3C,E-G). Comparing CMV-induced peripheral blood NK cells (cluster 6) to canonical CD56^dim^ NK cells (clusters 0 and 7), we observed the gene signature defined by Rebuffet *et al.* as NK3 cells and overlapping with previously described CMV-induced adaptive NK cells^20,21^, including genes *KLRC2, CD52, CCL5, GZMH, IL32, CD3E, VIM,* and *LGALS1* (Fig. 4A; Supplementary Table 9). Additional genes upregulated in our CMV-induced NK3 cells included *LAG3, PTMS, COL6A2, CRIP1, CD74* and members of the HLA-D gene family. In parallel, CMV-induced dNK cells (cluster 2) had a different set of genes upregulated compared to canonical dNK2/3 subsets (clusters 3 and 8), including *LAG3* and *HLA-DRB5* (Fig. 4B; Supplementary Table 10). Nine genes involved in NK cell activation (*FGFBP2, TNFRSF9, SPON2, FCGR3A, CCL4L2, S1PR5,* and *GZMB*), and one gene related to inhibition (*PTGDS*) made up the top upregulated genes in CMV-induced dNK cells (Fig. 4B). CMV-induced dNK cells expressed markers associated with tissue-residency, CD69 and CD49a/*ITGA1*, at both RNA and protein levels, indicating an additional fourth subset of dNK cells in the decidua of pregnant women with a history of CMV infection (Supplementary Fig. 6A). CMV-induced NK cells from peripheral blood and decidua lacked expression of *AREG* but it was upregulated in both non-CMV-induced CD56^dim^ NK and dNK2/3 cells (Fig. 4A,B). *AREG* (amphiregulin) is molecule known for its role in wound healing, and is associated with CD56^bright^/NK2 cells^13^. We observed that CMV-induced NK cells had lowest *AREG* expression across all NK cell clusters and groups, suggesting that there might be a link between CMV infection and the signalling pathways involved in *AREG* expression (Supplementary Fig. 6B,C)

**Figure 4.**
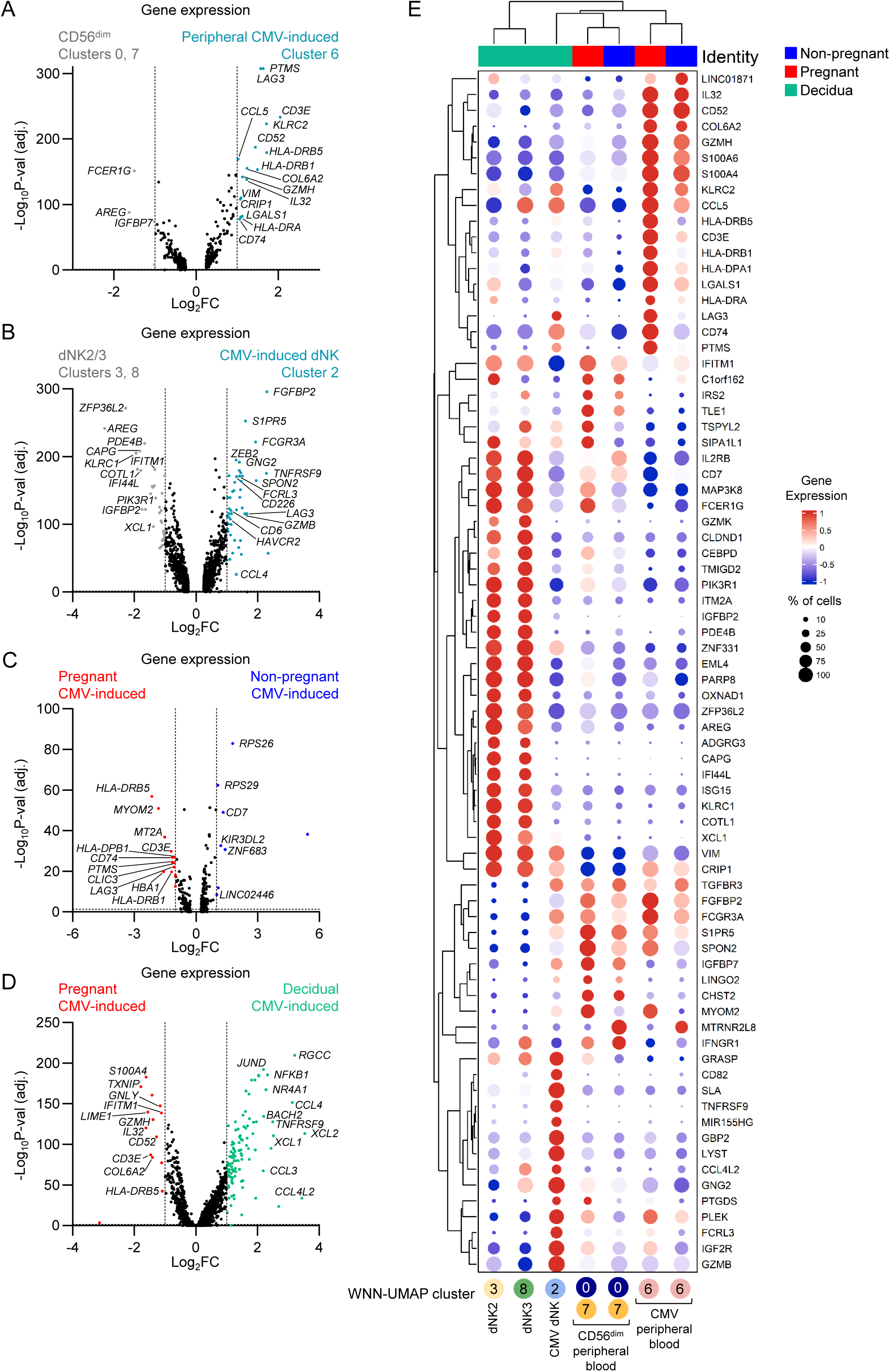
dNK4 subset in CMV-positive pregnancies. (A) Volcano plot of differential gene expression between peripheral CD56^dim^ (clusters 0 and 7) and CMV-induced CD56^dim^ (cluster 6) subsets. (B) Volcano plot of differential gene expression between dNK2 and 3 subsets (clusters 3 and 8) and CMV-induced dNK (cluster 2) subsets. (C) Volcano plot of differential gene expression between pregnant and non-pregnant CMV-induced NK cells (cluster 6). (D) Volcano plot of differential gene expression between pregnant and decidual CMV-induced NK cells (cluster 2). (E) Hierarchically clustered dot plot of the defining genes in CMV-induced NK cells. (A-D) Horizontal dashed lines depict statistical significance threshold, vertical dashed lines indicate the log_2_FC cutoff.

As CMV reactivation contributes to the global burden of pregnancy complications, with up to 42% of pregnant women developing non-primary CMV infections^22^, we sought to understand whether this CMV-induced gene signature is altered during pregnancy. When CMV-induced NK3 cells were compared between pregnant and non-pregnant women, we found higher expression levels of HLA-II related genes (*HLA-DRB5, HLA-DPB1, HLA-DRB1,* and *CD74*) and *LAG3* in pregnancy (Fig. 4C; Supplementary Table 11). We also examined differences between CMV-induced NK cells from pregnant peripheral blood and decidua. While pregnant peripheral blood CMV-induced NK cells had higher expression of the gene set associated with the NK3 subset (*CD52, GNLY, LIME1, CD3E, GZMH, IL32*), CMV-induced dNK cells had 109 genes upregulated by >2-fold (log_2_FC>1), the top 10 being *CCL4, XCL2, CCL4L2, RGCC, HSPA1A, XCL1, TNFRSF9, CRTAM, NR4A1,* and *NFKB1* (Fig. 4D; Supplementary Table 12). Similar to pregnant peripheral blood CMV-induced NK cells, CMV-induced dNK had high HLA-II gene and *CD74* expression (Fig. 4E; Supplementary Fig. 6D). Taken together, our data show that pregnancy affects the gene expression of CMV-induced NK cells, defined by increased HLA-II gene expression, *CD74*, and exhaustion marker *LAG3*. Additionally, we report that CMV-positive individuals have an additional NK cell subset within the decidua, which we named dNK4. Like other dNK subsets, dNK4 cells expressed tissue-residency markers CD69 and CD49a at the gene and protein levels, but otherwise had a unique gene set from peripheral CMV-induced NK cells and dNK2/3 subsets, with the most prominent features being *CD82, TNFRSF9, FCRL3, SLA, GBP2, IGFR2* (Supplementary Fig. 6E).

### **Pregnancy biases NK cells towards HLA-II,** *AREG* **and** *SOCS1* **expression**

To identify altered transcriptomic states of NK cells in pregnant peripheral blood a pseudo-bulk differential gene expression analysis was performed between pregnant and non-pregnant groups. This identified a total of 361 differentially expressed genes (DEGs), with 194 genes upregulated in pregnant NK cells (56 genes with >2-fold increase) and 167 genes upregulated in non-pregnant NK cells (41 genes with >2-fold increase) (Fig. 5A; Supplementary Table 13). Remarkably, 7 of the 56 genes (12.5%) that had >2-fold increase in pregnant NK cells encoded HLA-II related genes (members of the HLA-D gene family and *CD74*) (Fig. 5A,B). We confirmed upregulation of HLA-II associated genes at the protein level, with increased expression HLA-DR on pregnant peripheral blood and decidual NK cells (Fig. 5C). When HLA-II expression levels were assessed across clusters, as expected the highest expression was on CMV-induced NK cells (Fig. 4,5D). We then performed specific analysis of canonical (non-CMV-induced) CD56^dim^/NK1 cells (clusters 0 and 7) between pregnant and non-pregnant women (Fig. 5E). Indeed, even in CD56^dim^/NK1 subset, *HLA-DRB5* remained more highly expressed in pregnancy, indicating an association of increased HLA-II with pregnancy itself (Fig. 5E; Supplementary Fig. 6F; Supplementary Table 14). In addition to upregulation of HLA-DR, we also observed higher expression of proteins associated with lymphocyte regulation including CD25/IL-2RA, CD39, CD73, CCR5 and PD-1 on pregnant blood and decidual NK cells, while CX3CR1, KLRG1 and CD158 (KIR) were increased on non-pregnant peripheral blood NK cells (Fig. 5C).

**Figure 5.**
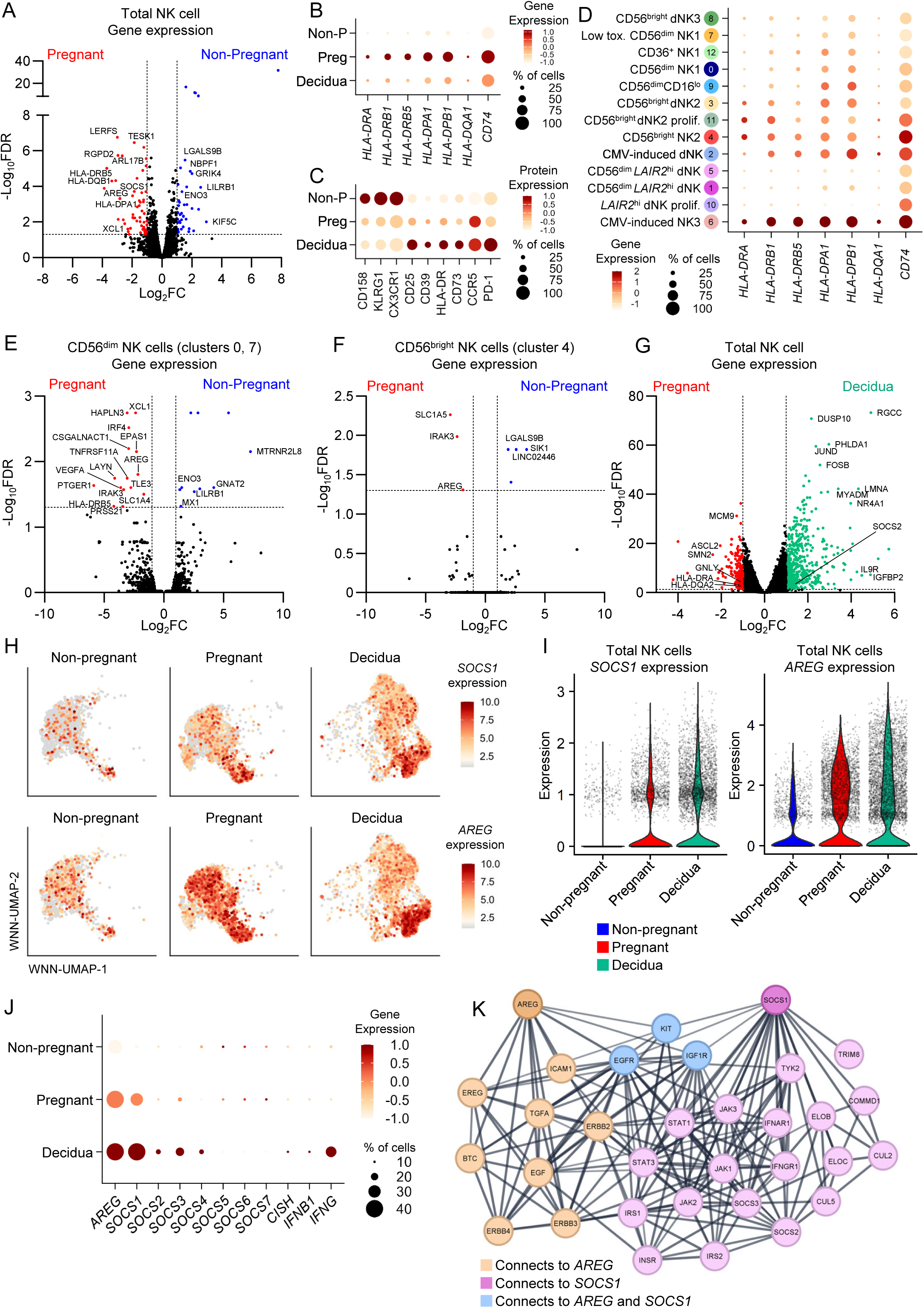
Identifying differentially expressed genes in NK cells during pregnancy. (A) Volcano plot of differential gene expression between pregnant and non-pregnant NK cells. (B,C) Bubble plot of expression level and proportion of cells expressing selected HLA-II associated genes (B) and selected differentially expressed proteins (C) per group. (D) Bubble plot of expression level and proportion of cells expressing selected HLA-II genes per WNN-UMAP cluster. (E,F) Volcano plots of differential gene expression between pregnant and non-pregnant (E) CD56^dim^ and (F) CD56^bright^ NK cells. (G) Volcano plot of differential gene expression between pregnant peripheral and dNK cells. (H) WNN-UMAPs split by group of *SOCS1* (top) or *AREG* (bottom) expression. (I) Violin plots showing *SOCS1* or *AREG* expression per group. (J) Bubble plot depicting *AREG*, *SOCS1-7, CISH, IFNB1* and *IFNG* expression level and proportion of cells expressing per group. (K) Protein-protein interaction network showing the top 30 interactions for a query of *AREG* and *SOCS1*. (B,C,D,J) Colour gradient depicts the gene/protein expression level. Circle size depicts the frequency of cells expressing. (A,E-G) Horizontal dashed lines depict statistical significance threshold, vertical dashed lines indicate the log_2_FC cutoff.

In addition to HLA-II associated genes, other key genes of interest upregulated in pregnancy were *SOCS1* (suppressor of cytokine signalling 1) and *AREG* due to their involvement in regulating immune responses (Fig. 5A). In fact, *AREG* was consistently upregulated across CD56^dim^/NK1 (clusters 0, 7) and CD56^bright^/NK2 (cluster 4) subsets in pregnancy, indicating a global increase in expression across all NK cells, although to a lesser magnitude in the CD56^bright^/NK2 subset (Fig. 5E,F; Supplementary Fig. 6G; Supplementary Tables 14-15). Given *AREG* is a defining feature of peripheral blood CD56^bright^/NK2 cells in a healthy state^13^, we reveal that higher *AREG* expression is observed in CD56^dim^/NK1 cells during pregnancy. Furthermore, expression of these key genes of interest (*HLA-D* family members, *SOCS1,* and *AREG*) in decidual NK cells as compared to pregnant peripheral blood was overall similar, with modestly higher expression of *SOCS1* (log_2_FC = 0.599) and *AREG* (log_2_FC = 0.197) in dNK cells, predominantly in the dNK2 (cluster 3) and dNK3 (cluster 8) subsets (Fig. 5G-I; Supplementary Table 16).

Suppressor of cytokine signalling (SOCS) proteins constitute a network of inhibitory molecules that regulate immune cell activation. We analysed expression levels of SOCS family member genes *SOCS1-7*, *CISH*, and cytokines *IFNB1* and *IFNG* to determine a pattern of expression for these related molecules. *SOCS1* had the highest expression level across all SOCS genes, especially in pregnant peripheral and dNK cells, while expression of other SOCS family member genes was relatively low (Fig. 5J). Within dNK cells, there was moderately higher expression of *SOCS2* (log_2_FC = 1.15), *SOCS3* (log_2_FC = 0.794) and *SOCS4* (log_2_FC = 0.608) as compared to pregnant peripheral NK cells (Fig. 5G,J; Supplementary Table 16). While amphiregulin and SOCS1 do not directly interact with each other, they both have direct interactions with KIT, EGFR and IGF1R, which are linked to JAK/STAT signalling pathways (Fig. 5K). This suggests that pregnancy increases expression of regulatory molecules AREG and SOCS1, as a potential mechanism to tightly control NK cell activation.

### **Higher gene signatures in** *AREG^+^ and SOCS1^+^* **NK cells during pregnancy**

With *AREG* typically associated with the CD56^bright^/NK2 subset, our observation that both CD56^bright^/NK2 cells and CD56^dim^/NK1 cells had higher *AREG* expression in pregnancy, led us to probe this subset further to understand its unique characteristics. Within peripheral blood, *AREG*^+^ NK cells had increased *FOS, COTL1, XCL1, GZMK, CD3E, IL7R, GRASP, SELL* and *XCL2,* many of which are associated the CD56^bright^/NK2 gene signature (Fig. 6A; Supplementary Table 17). Meanwhile, 43 genes were increased in *AREG^+^* dNK cells, with top hits including *XCL1, IGFBP2, IFI44L, ZFP36L2, CAPG, COTL1, KRT86, TNFRSF18, KLRC1* and *ZNF683* (Fig. 6B; Supplementary Table 18). Overall, there were more *AREG*^+^ NK cells in pregnant peripheral blood (73.9%) and decidua (64.5%) compared to non-pregnant blood (36.5%) (Fig. 6C). Gene signatures for *AREG*^+^ peripheral blood and decidual NK cells had four genes in common, namely, *XCL1, COTL1, XCL2* and *IL7R* (Fig. 6D,E). Many of the genes upregulated in peripheral blood *AREG*^+^ NK cells had higher expression during pregnancy (Fig. 6E). Further to this, CD56^dim^ NK cells from pregnant peripheral blood had higher expression of HLA-D family members, *JUND*, *GNLY*, *CCL5* and *CD3E*, with CD56^bright^ NK cells also associated with increased *HLA-DRB5* and *CD3E*, as well as *TXNIP* which has a suggested role in NK cell development and function^23^ (Supplementary Fig. 6H,I). Meanwhile, AREG^+^ CD56^dim^ and CD56^bright^ NK cells from non-pregnant blood had higher expression of ribosomal genes, potentially linked to enhanced NK cell function and activation.

**Figure 6.**
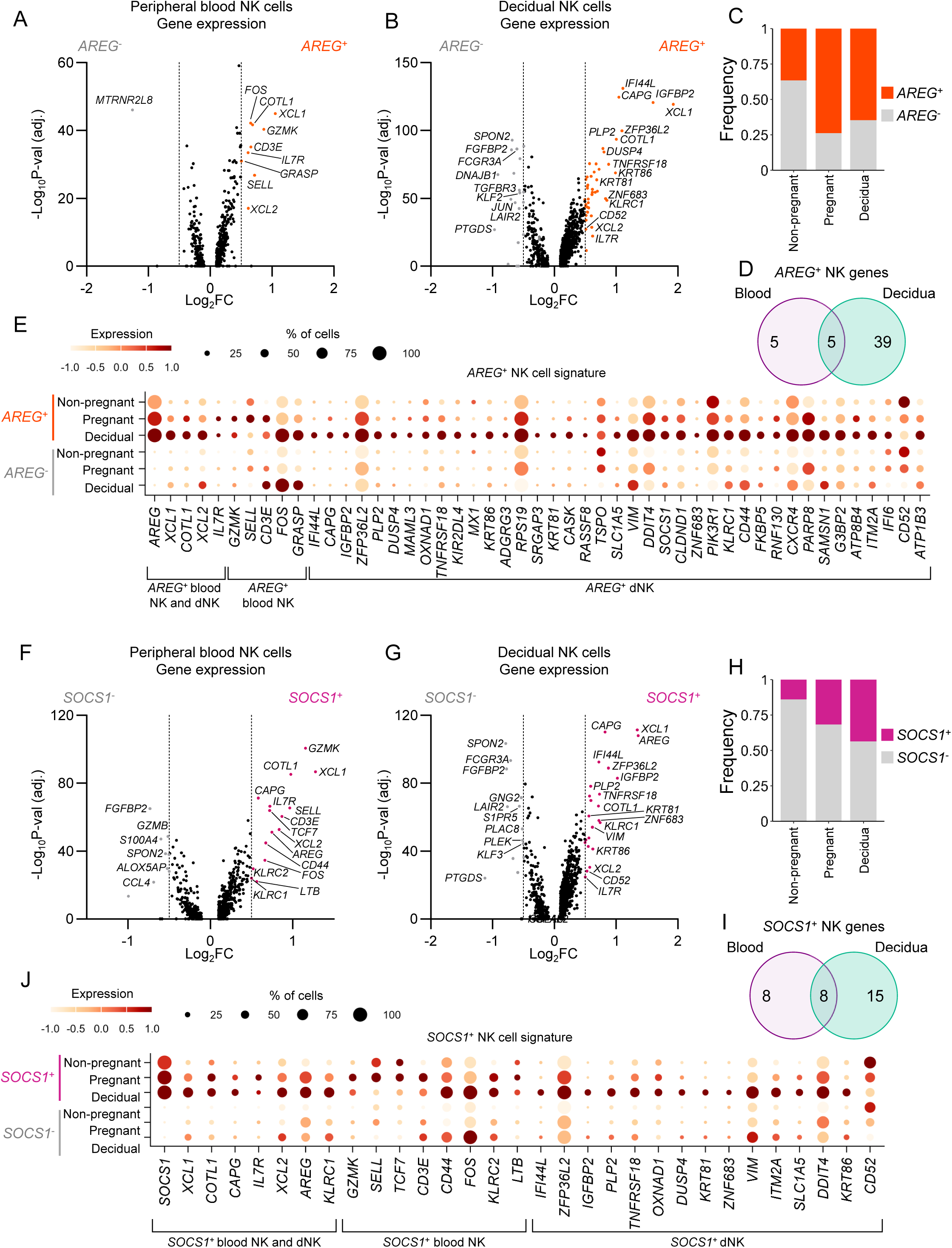
Defining features of *SOCS1*^+^ and *AREG*^+^ NK cells. (A-B) Volcano plots of differential gene expression between *AREG*^+^ and *AREG*^-^ NK cells from (A) peripheral blood and (B) decidua (excluding CMV-induced NK cells). (C) Frequency of *AREG*^+^ and *AREG*^-^ NK cells per group. (D) Venn diagram of genes expressed in *AREG*^+^ NK cells from peripheral blood and decidua. (E) Bubble plot of expression level and proportion of cells expressing genes upregulated in *AREG*^+^ NK cells across *AREG*^+^ and *AREG*^-^ NK cells per group. (F-G) Volcano plots of differential gene expression between *SOCS1*^+^ and *SOCS1*^-^ NK cells from (F) peripheral blood and (G) decidua (excluding CMV-induced NK cells). (H) Frequency of *SOCS1*^+^ and *SOCS1*^-^ NK cells per group. (I) Venn diagram of genes expressed in *SOCS1*^+^ NK cells from peripheral blood and decidua. (J) Bubble plot of expression level and proportion of cells expressing genes upregulated in *SOCS1*^+^ NK cells across *SOCS1*^+^ and *SOCS1*^-^ NK cells per group. (A,B,F,G) Horizontal dashed lines depict statistical significance threshold, vertical dashed lines indicate the log_2_FC cutoff.

There was no association of increased *SOCS1* within the *AREG*^+^ peripheral blood subset, however *SOCS1*^+^ NK cells had significantly higher expression of *AREG* than *SOCS1*^-^ NK cells (Fig. 6F,G; Supplementary Table 19,20). This indicated that *SOCS1*^+^ cells are *AREG*^+^, but there are *AREG*^+^ NK cells that do not express *SOCS1*, suggesting some level of differentiation associated with these markers. Pregnant peripheral blood and dNK cells had higher proportions of *SOCS1*^+^ NK cells (31.8% and 43.5%, respectively) than non-pregnant periphery (14.0%) (Fig. 6H). There were seven additional shared genes in *SOCS1*^+^ NK cells between peripheral blood and decidua, namely *XCL1, COTL1, CAPG, IL7R, XCL2, AREG* and *KLRC1*, of which *XCL1, XCL2, COTL1,* and *IL7R* were also associated with *AREG^+^* NK cells (Fig. 6I,J). Overall, we demonstrate that *AREG* is associated with the CD56^bright^NK2 subset and comprises a subset of *SOCS1*^+^ and *SOCS1*^-^ *AREG*^+^ NK cells. Our data provide important insights into NK cell subsets and their heterogeneity during pregnancy.

### Chemokine receptors define NK cell subset bias in pregnancy

Our high dimensional datasets revealed key differentially expressed genes and proteins on NK cells during pregnancy. We integrated these findings to define how pregnancy effects the expression of selected immune markers (CD6, CCR5, CD9, CD36, CX3CR1, CD39, CD73, HLA-DR, and Areg) on NK cells, and whether their expression can be modulated. Using additional samples, we analysed NK cells in whole blood from non-pregnant women, term pregnant women and decidua. Overall frequencies of CD6^+^, CX3CR1^+^, CD39^+^, CD73^+^ and Areg^+^ NK cells were comparable between pregnant and non-pregnant peripheral blood (Fig. 7A-C). Decidual NK cells, however, had lower frequencies of CD6^+^ and CX3CR1^+^ subsets but increased CD39, CD73 and Areg expression when compared to pregnant peripheral blood NK cells (Fig. 7A-C). While we identified higher *AREG* expression (36-74%) on NK cells at the

**Figure 7.**
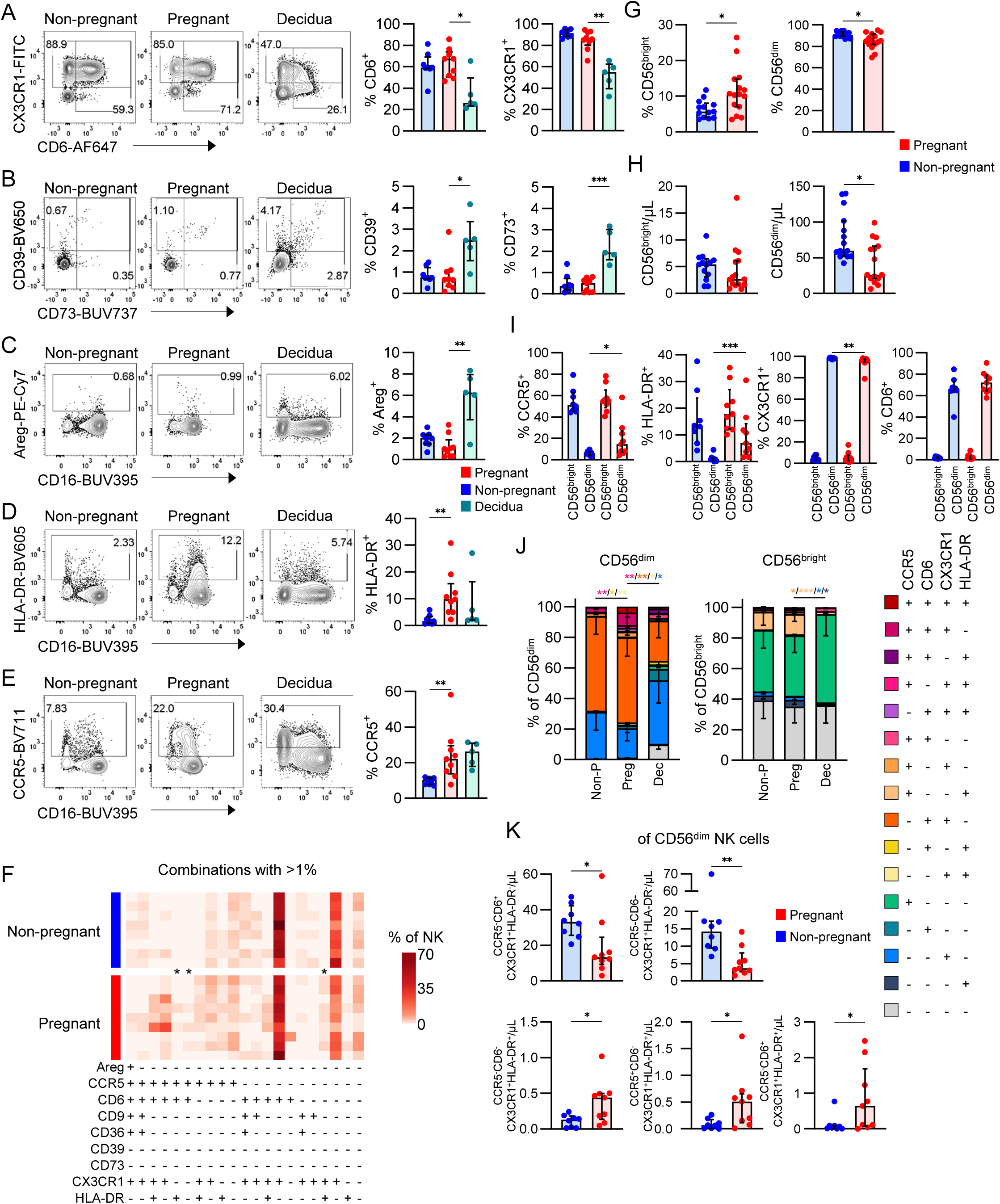
NK cell subset biases in pregnancy. (A-E) Representative FACS plots and frequencies of CX3CR1 and CD6 (A), CD39 and CD73 (B), Areg (C), HLA-DR (D), and CCR5 (E) in NK cells from pregnant and non-pregnant peripheral blood, and decidua. Statistics shown are a Mann-Whitney *U* test. (F) Heatmap depicting the frequencies of various combinations of Areg, CCR5, CD6, CD9, CD36, CD39, CD73, CX3CR1, and HLA-DR in pregnant and non-pregnant peripheral NK cells. (G) Frequencies of CD56^bright^ and CD56^dim^ NK cells in the pregnant and non-pregnant periphery. (H) Cell numbers of CD56^bright^ and CD56^dim^ NK cells per μL of whole blood in the pregnant and non-pregnant periphery. Statistics shown are a Mann-Whitney *U* test. (I) Frequencies of CCR5^+^, HLA-DR^+^, CX3CR1^+^, and CD6^+^ NK cells within the CD56^bright^ and CD56^dim^ subsets in the pregnant and non-pregnant periphery. (J) Frequencies of combinations of CCR5, CD6, CX3CR1, and HLA-DR in CD56^dim^ and CD56^bright^ NK cells from the pregnant and non-pregnant periphery, and decidua. (K) Numbers of set combinations of CCR5, CD6, CX3CR1, and HLA-DR in CD56^dim^ NK cells from pregnant and non-pregnant blood. All statistics shown are a Mann-Whitney *U* test, with the exception of (J) showing a two-way ANOVA.

RNA level in our scRNAseq dataset, low frequencies of Areg^+^ NK cells (<8%) were detected via flow cytometry, with dNK cells having higher frequencies than pregnant peripheral blood (Fig. 7C). We also confirmed via flow cytometry, higher frequencies of HLA-DR^+^ and CCR5^+^ NK cells in peripheral blood during pregnancy, as observed from our scRNAseq data (Fig. 7D-E). To determine whether these markers were co-expressed, we performed combinatorial analysis generating 512 unique combinations. Of these, 21 combinations had >1% frequency in NK cells which were predominantly defined by four markers, CCR5, CD6, CX3CR1 and HLA-DR. We identified three immune marker combinations that were higher in pregnant blood compared to non-pregnant blood, namely CCR5^+^CD6^+^, CCR5^+^CX3CR1^+^HLA-DR^+^ and CX3CR1^+^HLA-DR^+^ NK cells (Fig. 7F). CCR5 and CX3CR1 are both chemokine receptors known to mediate cell adhesion and migration, while CD6 is a co-stimulator scavenger receptor. In the context of NK cells, CD6 is expressed on a subset of peripheral blood CD56^dim^ NK cells, of which direct triggering can be linked to cytokine and chemokine secretion^24^.

We next determined whether CCR5, CD6, CX3CR1 or HLA-DR were associated with the CD56^dim^ or CD56^bright^ subsets. Firstly, while published literature suggests a shift towards a higher frequency of CD56^bright^ NK cells in the decidua^25,26^, literature is scarce regarding the makeup of CD56^dim^ or CD56^bright^ NK cell subsets in peripheral blood of term pregnancy^8^. Here, we found ∼2-fold increased frequencies of CD56^bright^ and decreased frequencies of CD56^dim^ subsets in pregnant blood compared to non-pregnant blood (Fig. 7G). Numbers of CD56^dim^ NK cells were also decreased, resulting in an increased proportion of CD56^bright^ and decreased proportion of CD56^dim^ cells (Fig. 7H). Of our key immune markers, CCR5 and HLA-DR were mainly enriched in CD56^bright^ NK cells in peripheral blood. Despite the association of these markers with CD56^bright^ cells, we observed higher frequencies of CCR5^+^ and HLA-DR^+^ CD56^dim^ NK cells in pregnant blood compared to non-pregnant blood, albeit at lower levels than CD56^bright^ NK cells (Fig. 7I). In contrast, CX3CR1 and CD6 were associated with CD56^dim^ NK cells, with modestly lower frequencies of CX3CR1^+^ CD56^dim^ NK cells in pregnant blood (Fig. 7I).

When we examined co-expression of the 4 key immune markers on CD56^bright^ and CD56^dim^ cell subsets, there were larger proportions of CD6^+^CX3CR1^+^ and CX3CR1^+^ cells within the CD56^dim^ subset, and CCR5^+^HLA-DR^+^ and CCR5^+^ within the CD56^bright^ subset (Supplementary Fig. 7A). In the pregnant group, CD56^dim^ cells had higher frequencies of CCR5^+^ CX3CR1^+^HLA-DR^+^, CD6^+^HLA-DR^+^, and CX3CR1^+^HLA-DR^+^ subsets, providing further evidence for increased HLA-II expression (Fig. 7J). Specifically, the reduced number of CD56^dim^ NK cells observed in pregnancy (Fig. 7H), could be attributed to lower numbers of CD6^+^CX3CR1^+^ and CX3CR1^+^ CD56^dim^ cells (Fig. 7K), indicating a global decrease in CX3CR1^+^ CD56^dim^ NK cells regardless of CD6 expression in pregnancy. Contrasting this, there were increased numbers of CX3CR1^+^HLA-DR^+^, CCR5^+^CX3CR1^+^HLA-DR^+^ and CD6^+^CX3CR1^+^HLA-DR^+^ NK cells within the CD56^dim^ subset in pregnancy (Fig. 7K, Supplementary Fig.), contributing to the observed increased frequency of HLA-DR^+^ CD56^dim^ NK cells (Fig. 7I). Across all combinations of CCR5, CD6, CX3CR1 and HLA-DR, we found no differences in cell numbers between pregnant and non-pregnant CD56^bright^ peripheral NK cells, further emphasizing that the CD56^bright^ subset is less affected by pregnancy (Supplementary Fig. 7C). These data suggest that pregnancy causes a skewing towards increased proportions of CD56^bright^ NK cells, through a numerical reduction in CX3CR1^+^CD56^dim^ populations.

### Type I inflammatory cytokines perturb the CX3CR1^+^ NK cell subset

Given that the NK cell subset bias in pregnancy was defined by a loss of CX3CR1^+^ CD56^dim^ cells, we investigated whether expression of CX3CR1, along with other differentially expressed markers, could be modulated by stimulation with soluble factors associated with type I (IL-12/IL-18 and IFNα) or type II (IL-17A, IL-33 and Areg) inflammation. CX3CR1 expression diminished in both pregnant and non-pregnant blood NK cells after stimulation with proinflammatory cytokines IL-12/IL-18, and to a lesser extent, IFNα (Fig. 8A,B). Increased frequencies of HLA-DR^+^ NK cells following IL-12/IL-18 stimulation were observed in both pregnant and non-pregnant blood (Fig. 8C,D). In contrast, CCR5 and CD6 expression did not change across stimulation conditions (Supplementary Fig. 8A-D). Both CX3CR1^+^CD6^+^ and CX3CR1^+^CD6^-^ populations decreased in frequency after IL-12/IL-18 stimulation, while the CX3CR1^+^CD6^+^ subset also decreased with IFNα stimulation (Supplementary Fig. 8E-F). This indicated the effects of IL-12/IL-18 and IFNα stimulation were specific for CX3CR1, given that CD6 expression did not change.

**Figure 8.**
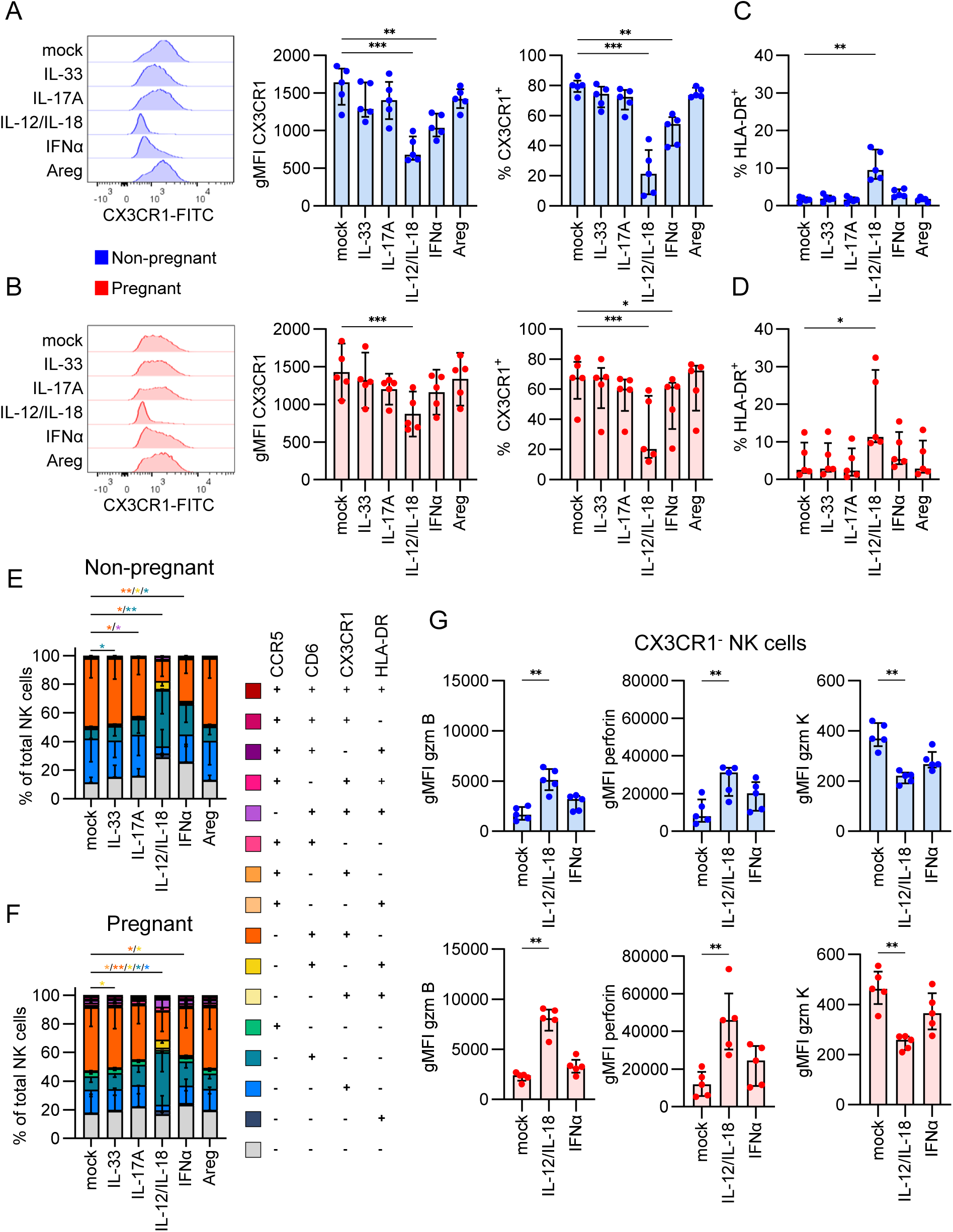
Modulation of CX3CR1 expression with type I cytokines. (A-B) Representative FACS histogram, gMFI, and frequency of CX3CR1 expression across mock, IL-33, IL-17A, IL-12/IL-18, IFNα, or Areg stimulation conditions in (A) non-pregnant and (B) pregnant NK cells. (C-D) Frequency of HLA-DR^+^ NK cells after mock, IL-33, IL-17A, IL-12/IL-18, IFNα, or Areg stimulation in (C) non-pregnant and (D) pregnant NK cells. (E-F) Proportions of NK cells with combinations of CCR5, CD6, CX3CR1, and HLA-DR in (E) non-pregnant and (F) pregnant NK cells. (G) gMFI of granzyme B, perforin, and granzyme K in CX3CR1^-^ and CX3CR1^+^ NK cells in non-pregnant and pregnant NK cells after mock, IL-12/IL-18, or IFNα stimulation. (A-D,G) Statistics shown are a Friedman test with Dunn’s multiple comparisons test. (E-F) Statistic is a two-way ANOVA.

We also assessed whether cytokine stimulation affected any combinations of CCR5, CD6, CX3CR1, and HLA-DR expression. Interestingly, we found a decrease in CD6^+^CX3CR1^+^ and increase in CD6^+^ NK cells in both pregnant and non-pregnant following IL-12/IL-18 stimulation, with less of an effect observed in non-pregnant blood following IFNα stimulation (Fig. 8E,F; Supplementary Table 21). In pregnant blood, the single-expressing CX3CR1^+^ NK cells were also decreased following IL-12/IL-18 stimulation further exemplifying the loss of CX3CR1 and maintenance of CD6 expression during IL-12/IL-18 and/or IFNα stimulation.

Given the evidence for a role of CX3CR1 in NK cell activation through IL-12/IL-18 and IFNα stimulation, we assessed functionality of CX3CR1^+^ and CX3CR1^-^ NK cell subsets by staining for granzymes, perforin, IFN-γ, TNF and MIP-1β (Supplementary Fig. 9A,B). Expression of granzyme B increased with IL-12/IL-18 stimulation in both CX3CR1^-^ and CX3CR1^+^ subsets (Fig. 8G left panel), but to a greater extent in the CX3CR1^-^ subset, for both pregnant (3.4-fold CX3CR1^-^, 1.5-fold CX3CR1^+^) and non-pregnant (3.1-fold CX3CR1^-^, 1.5-fold CX3CR1^+^) blood (Fig. 8G). Expression of perforin also increased while granzyme K decreased, but only in the CX3CR1^-^ NK cells, for both pregnant and non-pregnant blood (Fig. 8G middle and right panels). In line with CX3CR1^-^ cells being known cytokine producers^27^, we observed higher IFN-γ production in this subset after stimulation with IL-12/IL-18 (Supplementary Fig. 9C,D). Given that CX3CR1 expression diminished with IL-12/IL-18 stimulation, the increase in granzyme B and perforin and decrease in granzyme K observed in the CX3CR1^-^ subset may be explained by an increased proportion of cytotoxic CD56^dim^ cells within the CX3CR1^-^ population.

Overall, these data suggest a biological role for CX3CR1 following activation, potentially as a mechanism to prevent chemotaxis away from the site of inflammation, or that CX3CR1^+^ NK cells are more potent effectors and undergo activation-induced cell death. The deceased CX3CR1 expression following IL-12/IL-18 stimulation, reminiscent of type I inflammation, and the resulting activation profile within the remaining CX3CR1^-^ population, has mechanistic potential for the observed loss of CX3CR1^+^ CD56^dim^ NK cells during the third trimester of pregnancy where the immune system shifts from a predominantly anti-inflammatory Th2 state to a pro-inflammatory Th1 state.

## DISCUSSION

In this study, we identified key molecules involved in the altered immunological state of pregnancy using high-throughput cell surface proteome screening of >350 markers and scRNAseq with 130 CITE-seq antibodies. We discovered, at a phenotypic level, higher frequencies of NK cells expressing CCR5, HLA-DR, CD39, and CD73, and reduced frequencies of NK cells expressing CX3CR1, CD9, and CD36 in pregnancy. At the transcriptomic level, we revealed higher gene expression of regulatory molecules *AREG* and *SOCS1*, and HLA-D genes in pregnancy.

Higher HLA-II expression in NK cells from pregnant women was striking, given it was observed at both RNA and protein levels. The role of HLA-II on NK cells is not well defined or understood, therefore this finding provides insights into novel mechanisms of NK cell function. HLA-II molecules are known for their role in antigen presentation but are also described as activation markers for T and NK cells^7,28^. From a functional perspective, little is known about HLA-II^+^ NK cells. A mouse model showed that MHC-II^+^ NK cells resulted in reduced antigen-specific CD4^+^ T cell activation due to competitive antigen presentation between MHC-II^+^ NK cells, which lack costimulatory molecules CD80/CD86, and professional antigen-presenting dendritic cells^29^. While trogocytosis is one mechanism for NK cells to acquire HLA-II, and has also been shown in CD8^+^ T cells^30^, our findings indicate an increase in HLA-II at both transcriptional and protein levels, suggesting an intrinsic upregulation of HLA-II. This could be a potential regulatory mechanism to control CD4^+^ T cell activation in pregnancy, although future mechanistic studies are needed to confirm the significance of HLA-II^+^ NK cells, and what role they may have in pregnancy.

Migratory markers CX3CR1 and CCR5 were differentially expressed between NK cells from pregnant and non-pregnant women. We identified a strong association of CX3CR1 with CD56^dim^ cells and CCR5 with CD56^bright^ cells and showed that pregnant women had reduced numbers of CX3CR1^+^ CD56^dim^ cells in the blood. CX3CR1 has been shown to play an important role in NK cell adhesion and ability to lyse target cells^31–33^. Given that CX3CR1 expression was decreased in NK cells from term pregnant women, this suggests a potential regulatory role during late pregnancy. Additionally, we showed that CX3CR1^+^ and CX3CR1^-^ NK cells respond differently to cytokine stimulation, offering mechanistic insights as to why there are population shifts in pregnancy. In contrast, CCR5 was increased on NK cells from pregnant women. While the role of CCR5 on NK cells during pregnancy is not well-defined, NK cell migration to the lung during IAV infection is partially CCR5-dependent^34–36^. Therefore, an increase in CCR5^+^ NK cells during pregnancy could enhance NK cell migration to the lung during respiratory virus infections and offer protection, or alternatively, contribute to hyperactivation and disease severity. While numbers of CCR5^+^ NK cells were similar between pregnant and non-pregnant women, the higher proportion of the CCR5 subset out of total NK cells in pregnancy may influence immunity especially in the context of respiratory viral infections.

We identified differential expression of molecules involved in cell adhesion, including CD9 and CD36. Co-expression of CD9 and CD36 has been described on monocytes/macrophages^37^ and platelets^38^, while CD9 is known to be expressed on dNK cells^18^. Co-expression of CD9 and CD36 is not well described on peripheral blood NK cells. Our analyses showed that ∼15% of peripheral blood NK cells co-expressed CD9 and CD36, with a lower frequency (∼8%) observed during pregnancy. An *in vitro* model of myeloid progenitor cell differentiation into NK cells found that CD36 expression decreased, while CD56 increased^39^. Therefore, a potential reason for decreased CD36^+^ NK cells during pregnancy is the reduction in the CD56^dim^ subset. Another mechanism underpinning the presence of CD9 and CD36 on NK cells, and their reduced frequency during pregnancy is platelet aggregation. A study investigating immunity to COVID-19 found ICU patients had an increase in NK cell-platelet aggregates, based on the identification of cells expressing NK cell markers CD56 and CD16, and platelet markers CD62P and CD41^40^. There is a known increase in platelet-platelet aggregation during pregnancy^41,42^, which could affect the amount of platelets available for NK cell binding, resulting in the observed reduced frequency of CD9^+^CD36^+^ NK cells in pregnant women.

Expression of *AREG* and *SOCS1* were increased at the transcriptional level in NK cells from pregnant women, with *AREG* being consistently upregulated within both the CD56^bright^ and CD56^dim^ subsets. SOCS1 is a member of the suppressors of cytokine signalling, which are molecules induced by inflammation and have key roles in regulating immune responses (reviewed in ^43–45^). SOCS1 is known for inhibiting JAK-STAT signalling in response to type I (IFNα/β) and type II (IFN-γ) interferons (reviewed in ^45^). Amphiregulin is also known for having a regulatory role in immune responses, as it is strongly associated with CD4^+^ regulatory T cells and type II immunity^46,47^. Pregnancy is also characteristically biased towards type II immunity^1^, hence the increase in *AREG* could be linked to the tolerogenic state of pregnancy. Interestingly, we found that *AREG* expression was highest in dNK cells. Others have shown that AREG^+^ dNK are implicated in decidualization^48^. When we investigated whether SOCS1 and amphiregulin shared downstream signalling pathways, we identified molecules that led to JAK-STAT signal transduction, suggesting that it may be beneficial in pregnancy to mediate pathways that lead to increased inflammation.

Our analysis of term decidua identified that the defined CD39^+^KIR^+^ dNK1 subset is essentially absent at the end of the third pregnancy trimester, while others have shown high frequencies of dNK1 in decidua from early pregnancies^14^. A study analysing NK subsets in uterus transplant patients showed that all CD39^+^KIR^+^ NK cells in the recipient were recipient-derived, suggesting they are replenished from the periphery^18^. This suggests that later in pregnancy, there is less infiltration of NK cells to the placenta compared to early pregnancy, perhaps because the vasculature is already well established. Like us, others have shown dNK1 reduce with gestational age, providing an explanation for the absence of dNK1 cells in this full-term pregnancy dataset^15,49,50^.

Importantly, we revealed that in pregnancies with a history of CMV, there is a novel (previously not described) subset of dNK4 cells that have an adaptive, peripheral blood NK3-like gene signature, while also expressing tissue-residency markers CD69 and CD49d. CMV in pregnancy is of great concern, with 42% of pregnancies in CMV^+^ mothers experiencing a non-primary CMV infection^22^, and primary CMV infection in pregnancy resulting in vertical transmission rates of 30% in the first trimester and 72% in the third trimester^51^. Importantly, congenital CMV infection is linked to birth defects, including permanent vision impairments caused by chorioretinitis, hearing loss, cerebral palsy, seizures, and in rare cases, death^52,53^. CMV can directly infect cells in the decidua and cytotrophoblast^54^, therefore the presence of CMV-induced dNK cells at the maternal-fetal interface may play a protective role in controlling infection and preventing vertical transmission.

In summary, our novel high-dimensional datasets generated from this study provide key knowledge on the immunological landscape of pregnancy. It is important to consider the physiological state of pregnancy in understanding immunity towards infections and cancers, as the needs of immune cells often differ to non-pregnant individuals, yet pregnant women are often excluded from clinical trials due to safety concerns. Therefore, data provided here can potentially inform vaccine and therapeutic strategies, specifically geared towards managing infections, diseases and complications of pregnancy.

## Data Availability

All data produced in the present study are available upon reasonable request to the authors

## Acknowledgements

We thank Kaitlin Constable and Katelyn Dark for support with the pregnancy cohort and Jenny Anderson, the Melbourne Cytometry Platform (University of Melbourne) and the Flow Cytometry and Cell Sorting Shared Resource (St Jude Children’s Research Hospital) for technical assistance. This research was funded in whole or part by the National Health and Medical Research Council (NHMRC) Investigator Grants: EL1 to LCR (#2026357) and THON (#1194036), EL2 to DAW (#1174555) and L2 to KK (#2033783). For the purposes of open access, the authors have applied a CC BY public copyright licence to any Author Accepted Manuscript version arising from this submission. JRH is supported by the Melbourne Research Scholarship from The University of Melbourne. This work was supported by MRFF Award (#2016062) to KK, PGT, THON and LCR, and NIAID UO1 grant 1U01AI144616-01 “Dissection of Influenza Vaccination and Infection for Childhood Immunity” (DIVINCI) to PGT and KK.

## Author Contributions

KK led the study. KK and LCR supervised the study. JRH, THON, LCR and KK designed the experiments. JRH, LCR, THON, LFA, RRH, XJ, LK, EKA, AM, MP and PMS performed and analysed experiments. JRH, SL, IT, JS and JCC analysed data. PMS, AC, ME, CX, H-FK, MAAKK, DIG, LKM, ADB, SEN, AGB and PGT provided crucial reagents. NdA, KB, DAW, ML, SW and NJH recruited or provided patient cohorts. JRH, THON, LCR, and KK wrote the manuscript. All authors reviewed and approved the manuscript.

## METHODS

### Study approval and ethics statement

A total of 31 pregnant (31 samples) and 19 non-pregnant (27 samples) women, 9 decidual single-cell suspensions (8 pregnancy-matched), and 2 pregnancy-matched umbilical cord blood samples from a subset of pregnant women, were used in this analysis (Supplementary Table 22). The median age of pregnant and non-pregnant participants was 35 and 31 years, respectively. All pregnant blood samples were from full-term (>37 weeks) pregnancies. Experiments conformed to the Declaration of Helsinki Principles and the Australian National Health and Medical Research Council Code of Practice. Written informed consents were obtained from all donors prior to the study. The studies were approved by the Mercy Health (R14/25 and R04/29) and University of Melbourne (nos. 1443389, 2056901, 1443540, 2056761, 1955465, 2024-13344-58055-11, 2020-20782-12450-1) human research ethics committees (HRECS).

### Human blood processing

After obtaining written informed consent, whole blood was collected in heparinised blood tubes. Plasma was collected from whole blood in heparinised tubes by collecting supernatant after centrifugation (300x g, 10 min, 23°C). PBMCs were isolated from heparinised whole blood or buffy coats through density-gradient centrifugation (Ficoll-Paque, GE Healthcare, Uppsala, Sweden). Isolated PBMCs were cryopreserved in fetal calf serum with 10% DMSO.

### Human placenta tissue processing

Placenta processing was performed as described previously ^7^. Placentae were obtained within 15 min of birth and processed within 24 h of collection. Placental lobules (cotyledons) were obtained from multiple locations on the maternal surface, before the basal plate and chorionic surface were removed and villous tissue obtained from the middle cross-section. Briefly, placental tissue was washed in PBS and mechanically dissociated before being subjected to enzymatic digestion in RPMI 1640 medium supplemented with 2 mg/mL Collagenase D (Roche, Basel, Switzerland), 0.2 mg/mL DNAase I (Roche), 100 U/mL penicillin, 100 µg/mL streptomycin and 10 mM HEPES. Digested tissue was filtered and red blood cells lysed using 0.168 M NH_4_Cl, 0.01 mM EDTA, and 12 mM NaHCO3 in ddH_2_O. Single-cell suspensions of decidual tissue were cryopreserved as described above for human blood processing.

### Cytomegalovirus (CMV) serology

Serology was performed to determine the CMV infection status of participants. Blood plasma was subjected to a LIASON® CMV IgG II assay (DiaSorin, Italy) to detect CMV-IgG using a chemiluminescent immunoassay.

### Flow cytometry on fresh whole blood

The applicable antibody mix (Supplementary Tables 7 and 23) was added directly to 200 µL of whole blood and incubated for 30 min at ambient temperature. Fluorescently-labelled samples were kept in the dark throughout the staining process. Red blood cells were lysed with 1X BD FACS™ Lysing solution (cat. no. 349202; diluted 1:10 with sterile H_2_O (Baxter, NSW, Australia)) for 10 min at ambient temperature. Samples were centrifuged (500 *g*, 5 min) and supernatant aspirated. Samples without required intracellular staining were resuspended in 1% PFA for 20 min at 4°C, before washing and resuspending in MACS buffer for acquisition on a BD LSRFortessa II. Samples requiring intracellular staining were permeabilized with the eBioscience^TM^ Foxp3/Transcription Factor Staining Buffer Set (Thermo Fisher Scientific; cat. no. 00-5523-00) for 20 min at 4°C before washing and resuspended with intracellular antibody mix made with the Permeabilization Buffer for a further 30 min at 4°C. Intracellular stain was washed with Permeabilization Buffer and samples were resuspended in MACS buffer for acquisition on a BD LSRFortessa II.

### Surface proteome screening with LEGENDScreen

Human LEGENDScreen PE kit (Biolegend, cat. no. 700011) antibodies (Supplementary Table 24) were reconstituted according to the manufacturer’s protocol. All antibody incubations were performed for 30 min, on ice and in the dark unless otherwise stated. PBMCs and placenta single-cell suspension samples were thawed with Benzonase nuclease (Merck Life Science Pty Ltd cat. no. E8263-25KU) before staining in 6×10^6^ cell aliquots for optimal labelling. Individual samples were barcoded with combinations of Cell Trace Violet (CTV) and/or anti-CD45-biotin plus secondary labelling with fluorochrome-conjugated streptavidin to allow pooling of donors for immune analysis, adapted from^55^. Samples requiring CTV labelling were resuspended in 1 µM CTV (ThermoFisher Scientific, VIC, Australia) for 13 min at 37°C (gentle vortexing every 3-4 min). CTV-labelled samples were washed, and all samples stained with 1:100 anti-CD45-biotin (Biolegend). Samples were washed twice in MACS buffer (0.5% BSA, 2mM EDTA in PBS) and resuspended in donor-specific barcode mixes containing streptavidin-BUV395, -BUV496, and/or -BUV805 (Supplementary Table 1). Following washing (x2), samples were stained with MR1-5-OP-RU-BV421 tetramer (1:100) for 30 min at room temperature. Samples were washed twice and resuspended in the backbone panel to determine immune cell populations (Supplementary Table 2). Samples were washed, pooled and evenly distributed across the Human LEGENDScreen PE kit, where cells were stained per the manufacturer’s protocol. Samples were resuspended in MACS buffer and acquired on the Cytek Aurora 5-laser flow cytometer.

### Surface proteome data analysis

LEGENDScreen flow cytometry data were analysed by first deconvoluting individual samples with donor barcode fluorophores. Individual sample gates were exported as new .fcs files. Manual gating was performed on individual marker-PE wells to determine frequencies of marker^+^ cells for each immune cell subset. LEGENDScreen markers overlapping with the backbone panel were not included in differential expression analysis due to potential competitive binding, and only markers with a frequency of ≥ 10% in at least one sample were analysed for frequencies. To reduce the number of false-positive hits, we utilised a combination of statistical significance, a log_2_ fold-change (log_2_FC) > 0.5 and a requirement of 20% of cells expressing in at least one group per comparison.

### scRNAseq of PBMCs and decidual NK cells

PBMCs and decidual single-cell suspension samples were thawed, washed and resuspended in 50 µL of MACS buffer. Samples were sequential stained at 4°C and in the dark with 5 µL of TruStain FcX (Biolegend, cat. no. 422301) incubated for 10 min; fluorescent monoclonal antibody mix (Supplementary Table 25) for 30 min; 20 µL of TotalSeq™-C Human Universal Cocktail (Biolegend, cat. no. 399905) for 30 min. Samples were washed, resuspended with MACS buffer before NK cells were sorted into RPMI with 10% fetal calf serum on a Sony SY3200 cell sorter (Sony Biotechnology, Illinois, USA). Sorted cells were counted and mixed into 10x Chromium reactions before being loaded into Chromium Next GEM Chip K (PN 2000182, CA, USA) and run on the Chromium Controller iX according to manufacturer’s instructions (Chromium Next GEM Single Cell 5’ v2 (Dual Index) with Feature Barcoding technology; Revision D). Gene expression (GEX) and feature barcode (CITE-seq) libraries were generated with Chromium Next GEM Single-Cell 5′ v2 kits according to manufacturer’s instructions. Sequencing was performed using an Illumina NovaSeq at a read length of 26 x 90 bp.

### scRNAseq data analysis

Raw sequencing data were processed with Cell Ranger v7.1.0 (10x Genomics). Donors were demultiplexed based on differences in single nucleotide polymorphisms using cellsnp-lite v1.2.2 and vireo v0.2.3^56,57^. Gene expression (GEX) and Feature Barcode (CITE-seq) matrices were analysed with the Seurat package v4.3.0.1^58^. Gene expression data were filtered for high-quality cells by removing cells with more than 10% mitochondrial mRNA, and cells with <500 or >5000 genes. CITE-seq data were filtered by removing cells with total reads outside 3× the median absolute deviation from the median library size. Sample demultiplexing was performed using the hashedDrops function from DropletUtils^59^. Weighted-nearest-neighbour (WNN) Uniform Manifold Approximation Projection (UMAP) was performed to generate cell clusters using integrated gene and protein level data. The RunHarmony function from the Harmony R package^60^ was applied to RNA and CITE-seq level data for donor/batch integration before running a WNN-UMAP. The FindMarkers function from Seurat was used to identify differentially expressed genes (DEGs) and proteins (DEPs) between clusters to help in assigning them to a relevant NK cell subset. Group analysis between pregnant and non-pregnant, or pregnant peripheral and placental NK cells was performed using a pseudo-bulk approach after removing unwanted variance (RUV). Pseudo bulking was performed by aggregating counts across groups, donors, and Harmony clusters. Unwanted variation was estimated with RUV-4 algorithm implemented in the ruv R package^61^ with k=4. Differential expression (DE) analysis was performed using the edgeR package^62^. Data were fitted with a negative binomial generalized log-linear model implemented in glmFit function^63^. Genes were considered differentially expressed between groups if they achieved an FDR <5% after likelihood ratio tests. DE analysis of CMV-induced subsets, and *SOCS1* and *AREG* subsets was performed using the Seurat FindMarkers function.

### Protein-protein interaction network

Protein-protein interaction analysis and network was generated in Cytoscape^64^ using the STRING *Homo sapiens* database. The genes of interest (*AREG* and *SOCS1*) were queried with a maximum additional interactor of 30, and a confidence score cutoff of 0.7.

### *Ex vivo* **PBMC stimulation with cytokines/soluble factors**

Cryopreserved PBMCs from pregnant and non-pregnant women were thawed and washed in cRPMI. Cells were resuspended in cRPMI at a concentration of 3×10^6^ cells/mL and 200 μL per condition were distributed into a 96-well U-bottom plate. PBMCs were stimulated with IFN-α (5000 U/mL), IL-33 (20 ng/mL), Amphiregulin (100 ng/mL), or IL-12 (10 ng/mL) and IL-18 (100 ng/mL) for 48 h. In the final 16 h of stimulation, brefeldin A was added (GolgiPlug, BD Biosciences cat. no. 555029) to samples for intracellular staining of IFN-γ, TNF, and MIP-1β. At the 48 h timepoint, supernatants were collected from the samples that did not receive brefeldin A, for use in the LEGENDplex assay (described below). Surface and intracellular staining was performed by washing samples with MACS buffer and adding 50 μL of applicable surface stain. Sample were washed before performing intracellular staining with the applicable panel as described in *Flow cytometry on fresh whole blood* (Supplementary Tables 26-27).

---

**Supplementary Figure 1.**
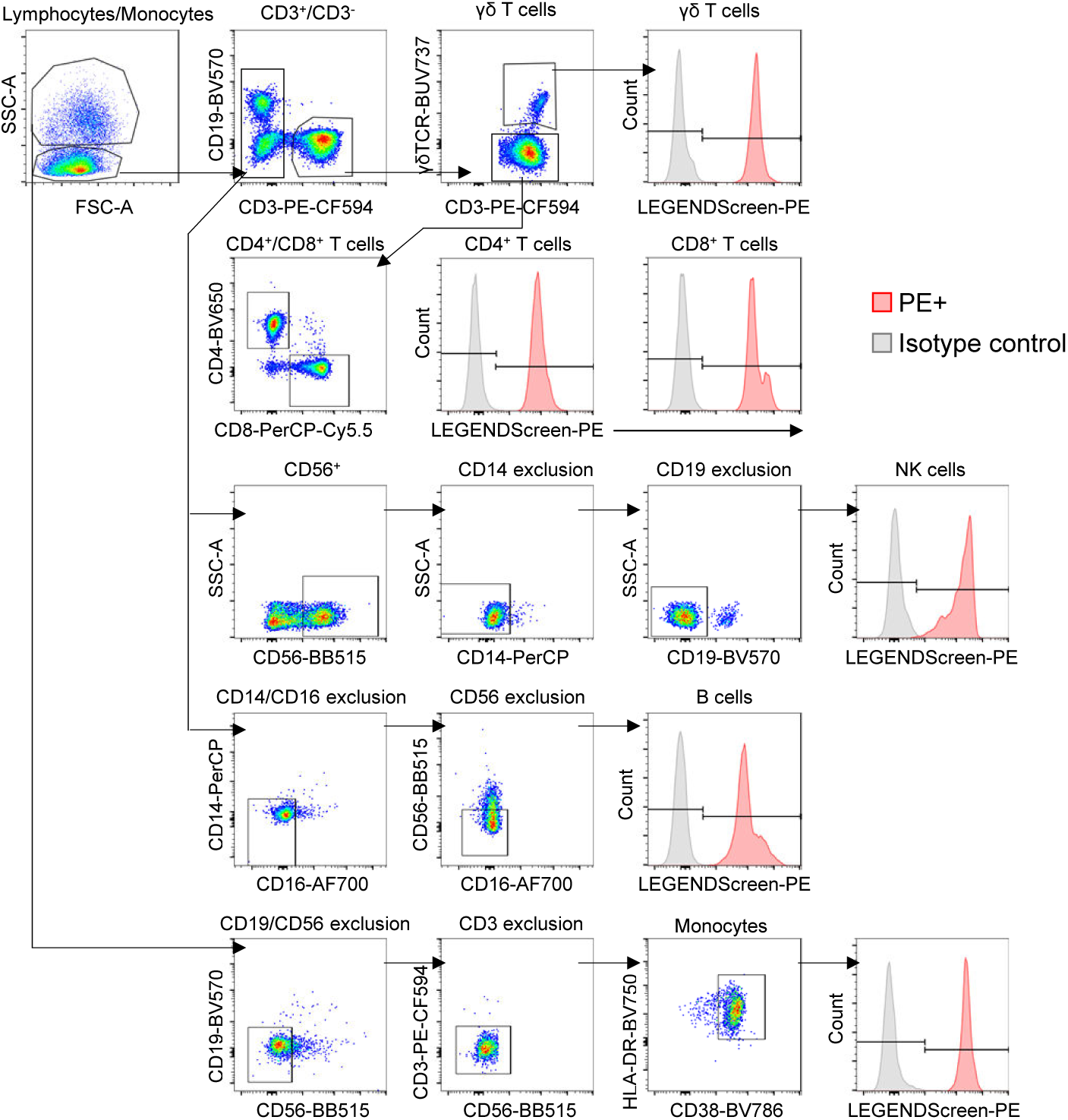
Gating strategy to identify NK cells, monocytes, B cells, CD8^+^ T cells, CD4^+^ T cells, and γδ T cells. Gating of LEGENDScreen mAb-PE signals (red) on histograms was based on respective isotype control (grey).

**Supplementary Figure 2.**
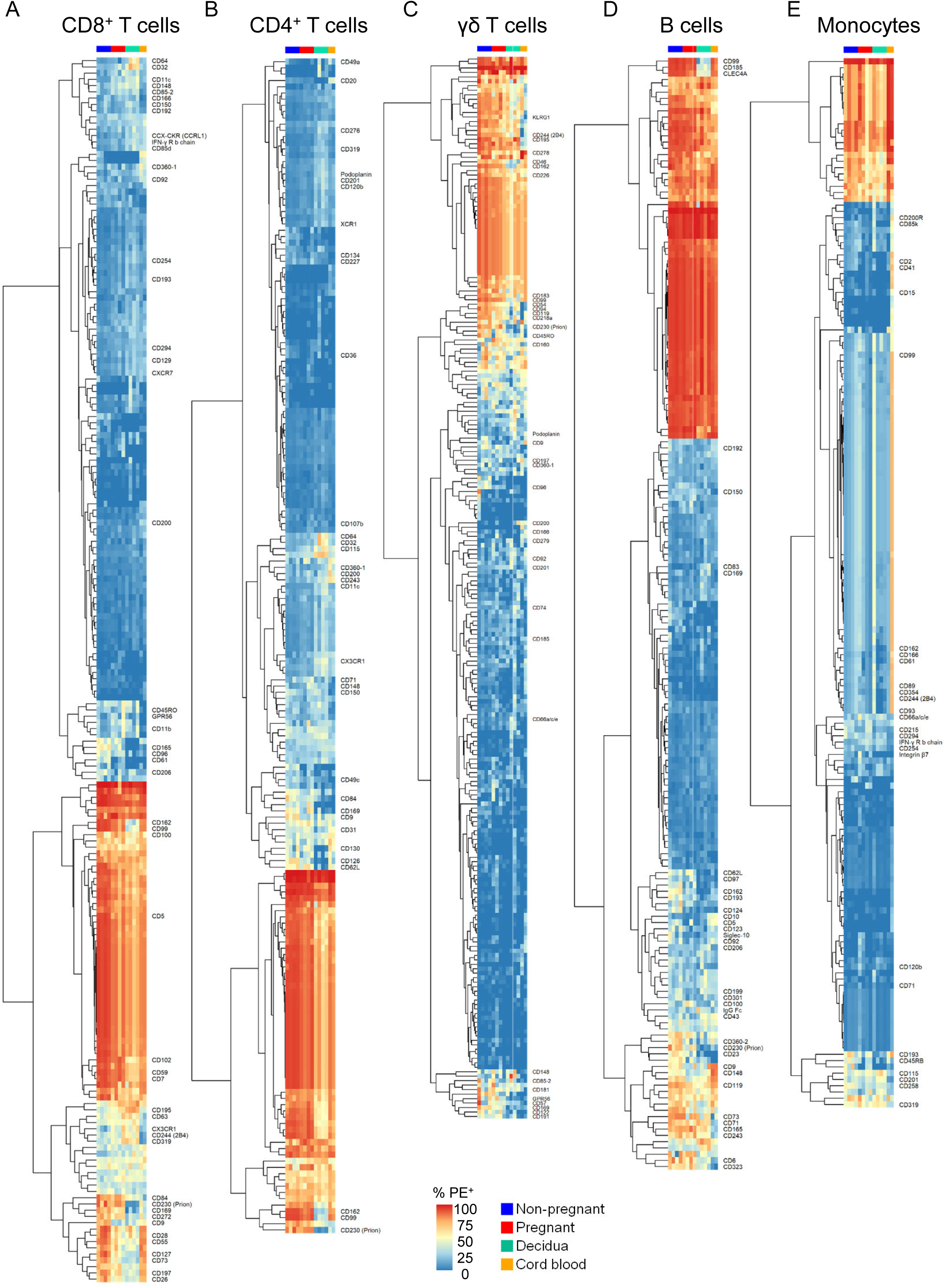
(A-E) Heatmaps with hierarchical clustering of columns showing frequencies of marker-PE expression on CD8^+^ T cells (A), CD4^+^ T cells (B), γδ T cells (C), B cells (D), and monocytes (E) from pregnant and non-pregnant peripheral blood, cord blood and decidua. Markers expressed on >10% of cells in at least one sample were included for analysis (196 (A), 188 (B), 238 (C), 178 (D), and 168 (E) out of total 342 markers).

**Supplementary Figure 3.**
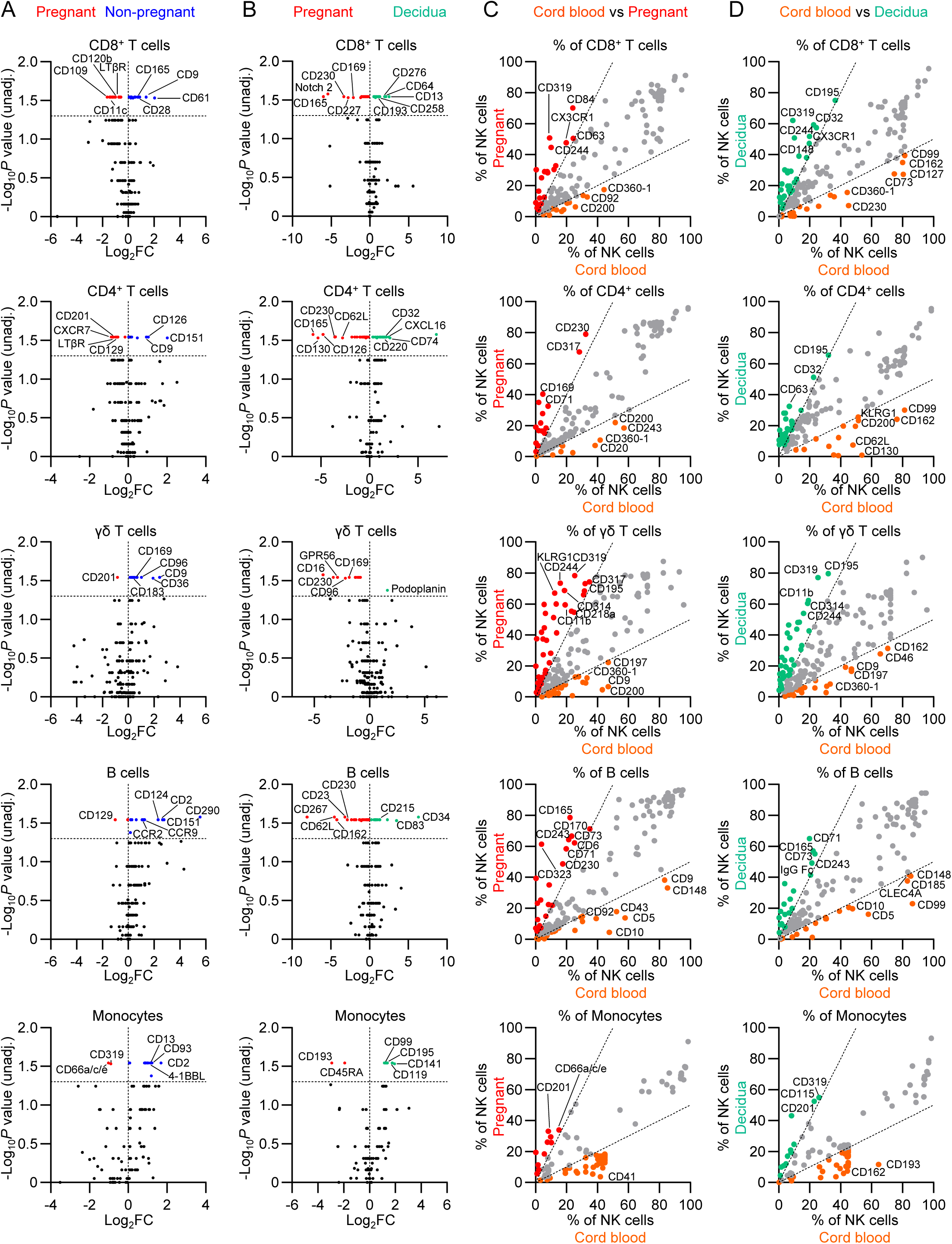
(A) Volcano plots depicting differential protein expression in non-pregnant vs pregnant peripheral blood immune cell populations including, CD8^+^ T cells, CD4^+^ T cells, γδ T cells, B cells and monocytes. (B) Volcano plots depicting differential protein expression in decidual vs pregnant blood immune cell populations. Volcano plot horizontal dashed line indicates the log_10_ p-value cutoff of 1.3 (p=0.05). (C,D) Scatter plot of average percent of cells expressing for (C) cord blood vs pregnant peripheral blood and (D) cord blood vs decidual CD8^+^ T cells, CD4^+^ T cells, γδ T cells, B cells, monocytes. Scatter plot dashed lines indicate a log_2_FC cut-off of 1.

**Supplementary Figure 4.**
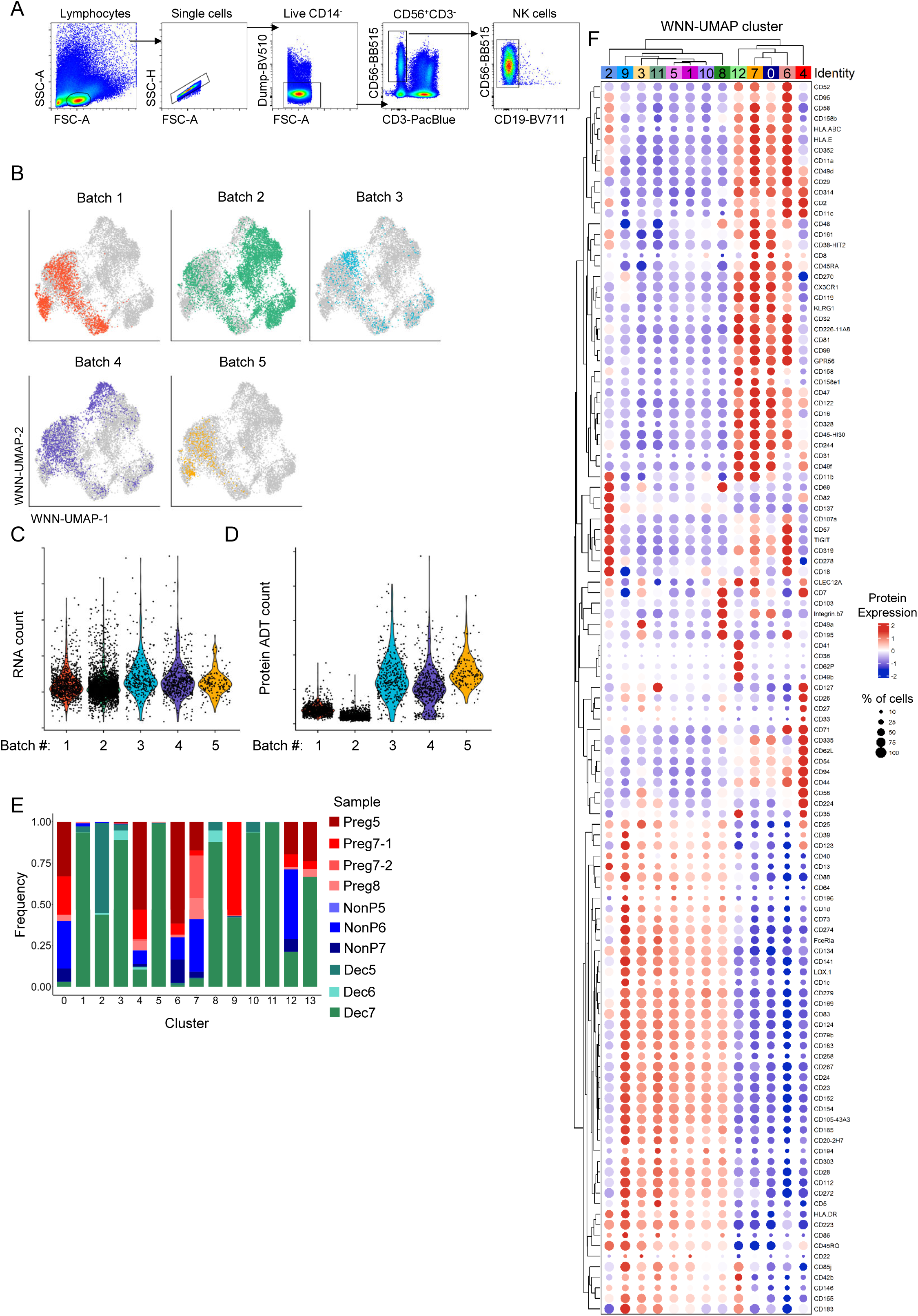
Sort gating strategy and quality control analysis of scRNAseq. (A) Gating strategy for bulk NK cells. (B) WNN-UMAP coloured by 10x reaction batches. (C) RNA and (D) antibody-derived-tag (ADT; C) read counts per cell, per batch of 10x reactions. (F) Proportion of each sample (pregnant peripheral blood NK cells (reds), non-pregnant peripheral blood NK cells (blues), decidua NK cells (teals) per WNN-UMAP cluster. (G) Hierarchically clustered dot plot of CITE-seq markers across each WNN-UMAP cluster.

**Supplementary Figure 5.**
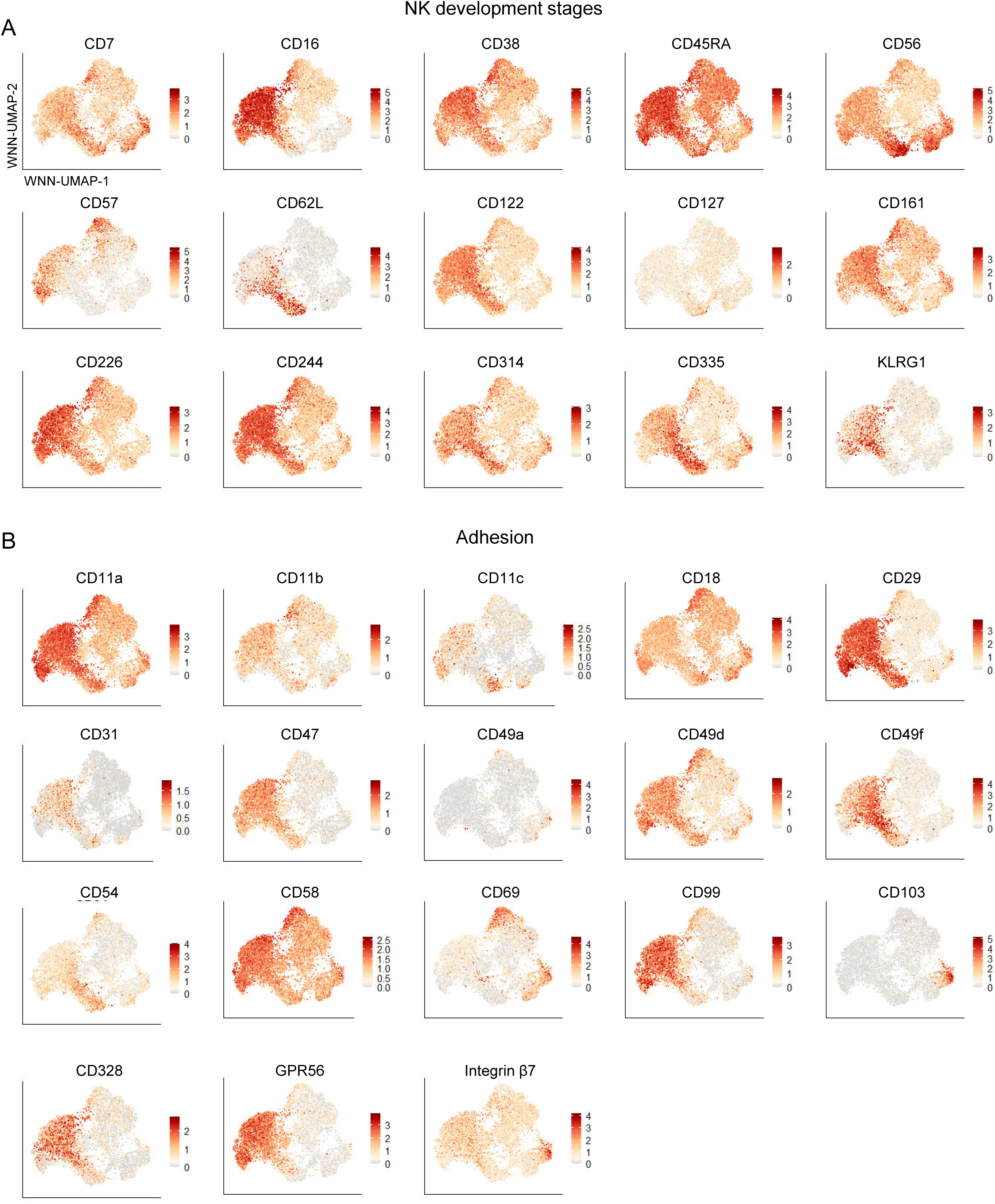
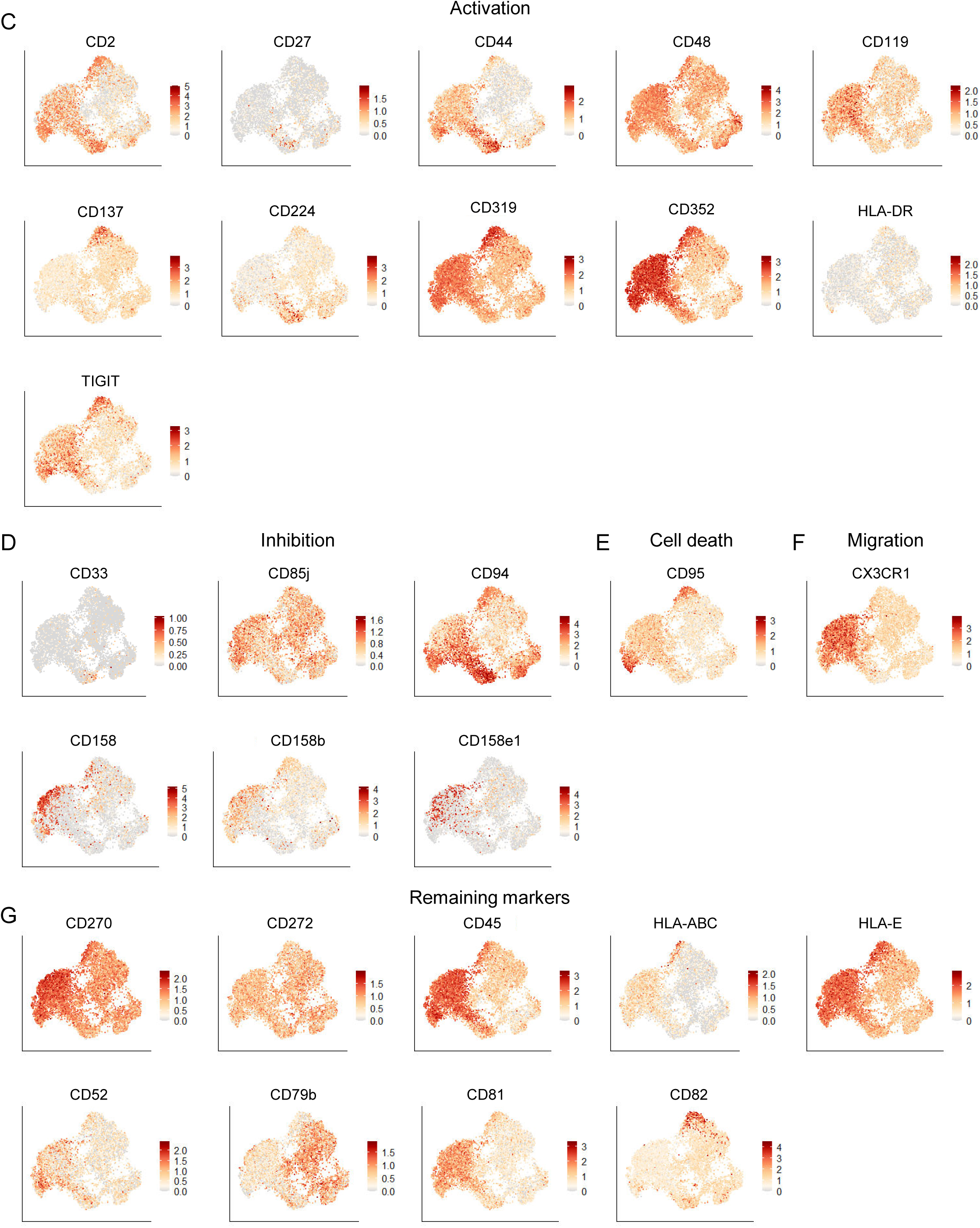
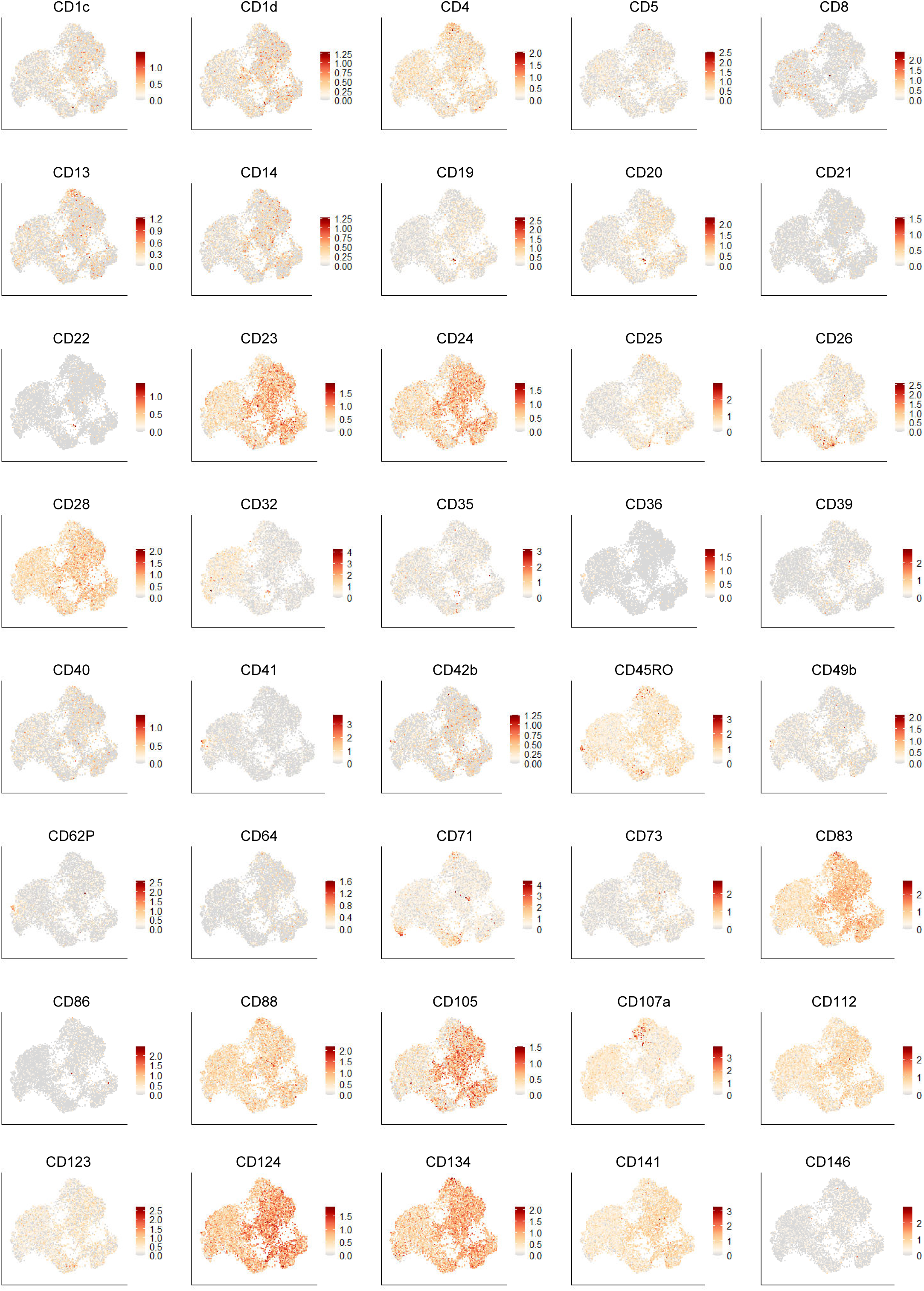
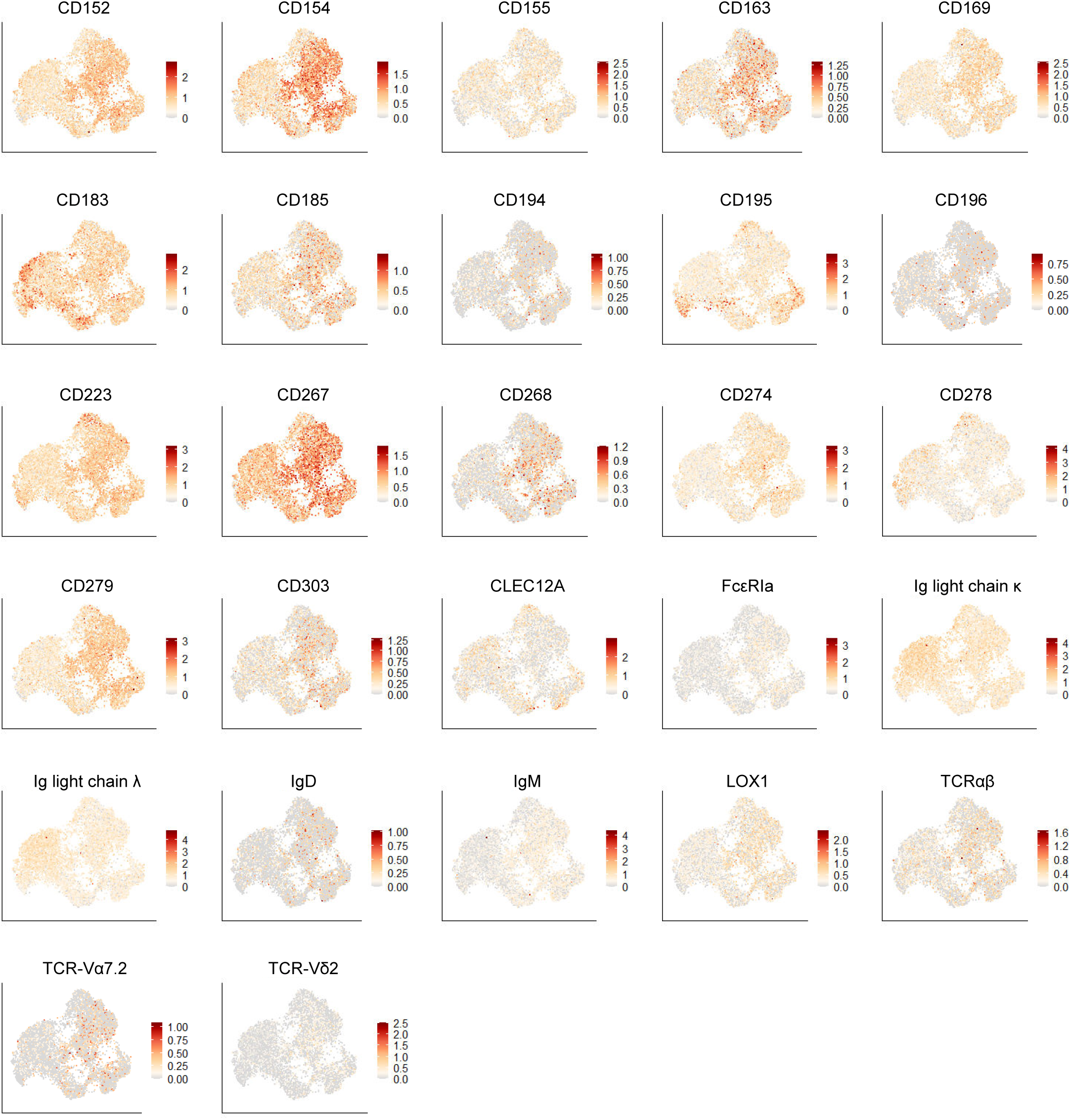
WNN-UMAPs depicting each of the 130 surface proteins detected with CITE-seq oligo-labelled mAbs. WNN-UMAPs are separated by proteins associated with (A) NK developmental stages, (B) adhesion, (C) activation, (D) inhibition, (E) cell death, (F) migration, and (G) remaining markers.

**Supplementary Figure 6.**
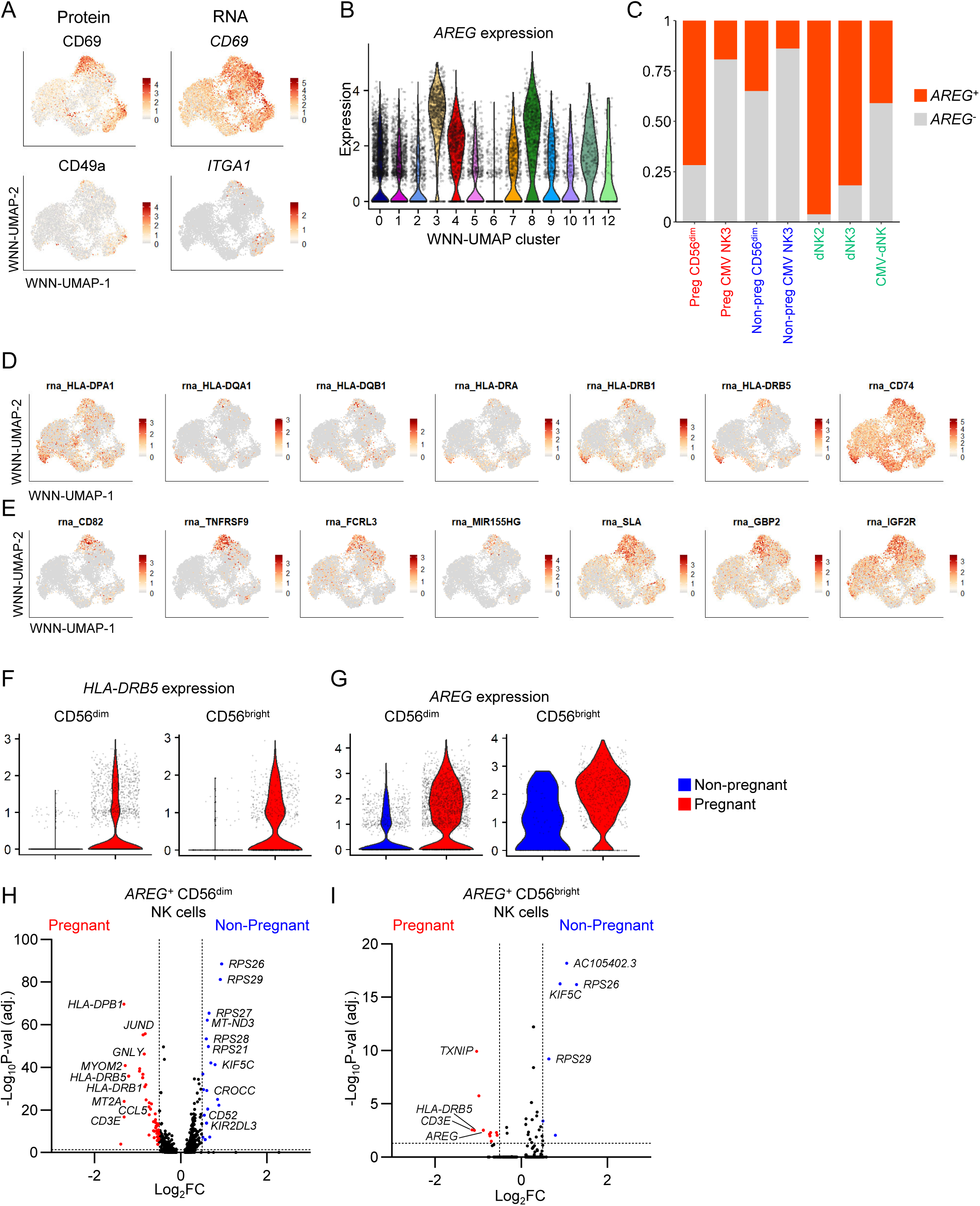
Markers associated with CMV-induced NK cells and pregnancy. (A) WNN-UMAPs showing expression of CD69 and CD49a/ITGA1 at the protein and RNA levels. (B) Violin plot depicting *AREG* expression across WNN-UMAP clusters. (C) Proportion plot showing the frequencies of *AREG*^+^ and *AREG^-^* NK cells across pregnant and non-pregnant peripheral blood non-CMV-induced CD56^dim^ NK cells and CMV-induced CD56^bright^ NK3 cells, and dNK2/3 and CMV-induced dNK cells. (D,E) WNN-UMAPS showing expression of (D) HLA-II genes and *CD74* and (E) selected genes associated with CMV-induced dNK cells (E). (F,G) Violin plot of (F) *HLA-DRB5* and (G) *AREG* expression in CD56^dim^/NK1 and CD56^bright^/NK3 cells from pregnant and non-pregnant peripheral blood. (H,I) Volcano plot of differential gene expression between pregnant and non-pregnant *AREG*^+^ CD56^dim^ (H) and *AREG*^+^ CD56^bright^ (I) NK cells.

**Supplementary Figure 7.**
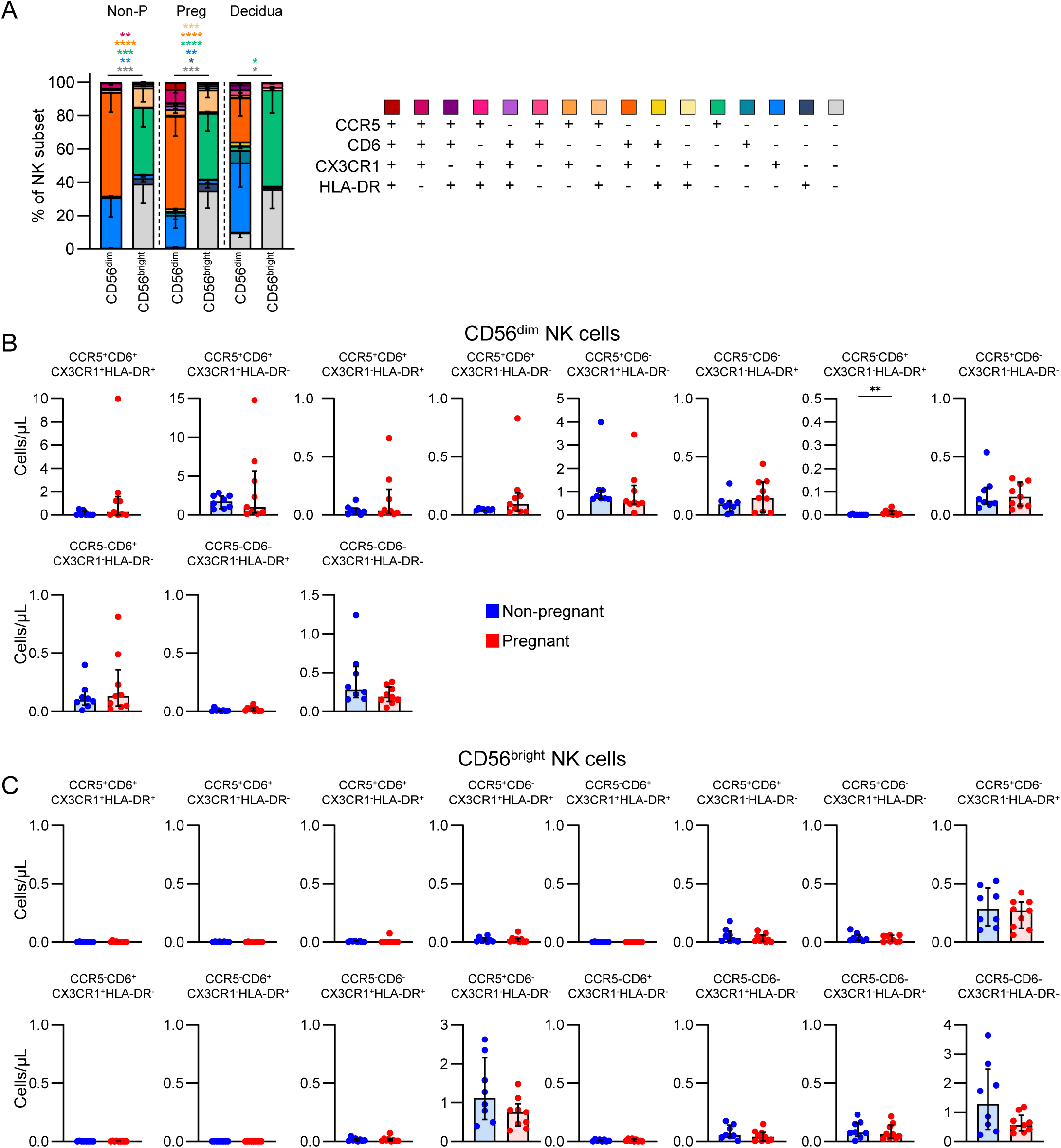
Numbers and frequencies of CCR5, CD6, CX3CR1, and HLA-DR combinations. (A) Frequencies of combinations of CCR5, CD6, CX3CR1, and HLA-DR in CD56^dim^ and CD56^bright^ NK cells from the pregnant and non-pregnant periphery, and decidua. Statistic shown is a 2-way ANOVA performed within each group. (B,C) Numbers of CCR5^+^, HLA-DR^+^, CX3CR1^+^, and CD6^+^ NK cells within the CD56^dim^ (B) and CD56^bright^ (C) subsets in the pregnant and non-pregnant blood. Statistics shown are a Mann-Whitney *U* test.

**Supplementary Figure 8.**
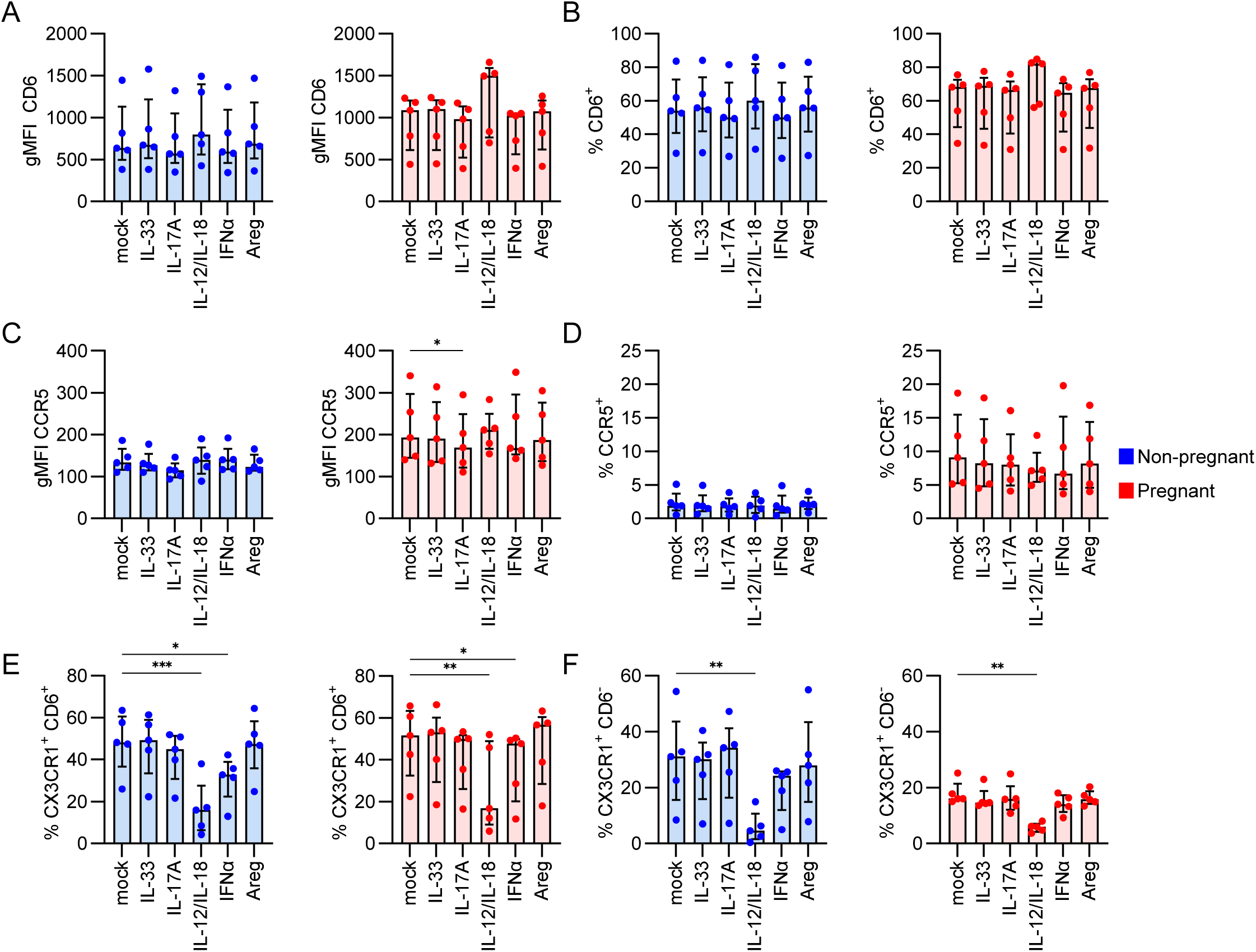
Phenotype shifts with cytokine stimulation. (A) gMFI and (B) frequency of CD6 expression across mock, IL-33, IL-17A, IL-12/IL-18, IFNα, or Areg stimulation conditions in non-pregnant and pregnant NK cells. (C) gMFI and (D) frequency of CCR5 expression across mock, IL-33, IL-17A, IL-12/IL-18, IFNα, or Areg stimulation conditions in non-pregnant and pregnant NK cells. (E,F) Frequency of (E) CX3CR1^+^CD6^+^ and (F) CX3CR1^+^CD6^-^ NK cells across mock, IL-33, IL-17A, IL-12/IL-18, IFNα, or Areg stimulation conditions in non-pregnant and pregnant groups. Statistics shown are a Friedman test with Dunn’s multiple comparisons test.

**Supplementary Figure 9.**
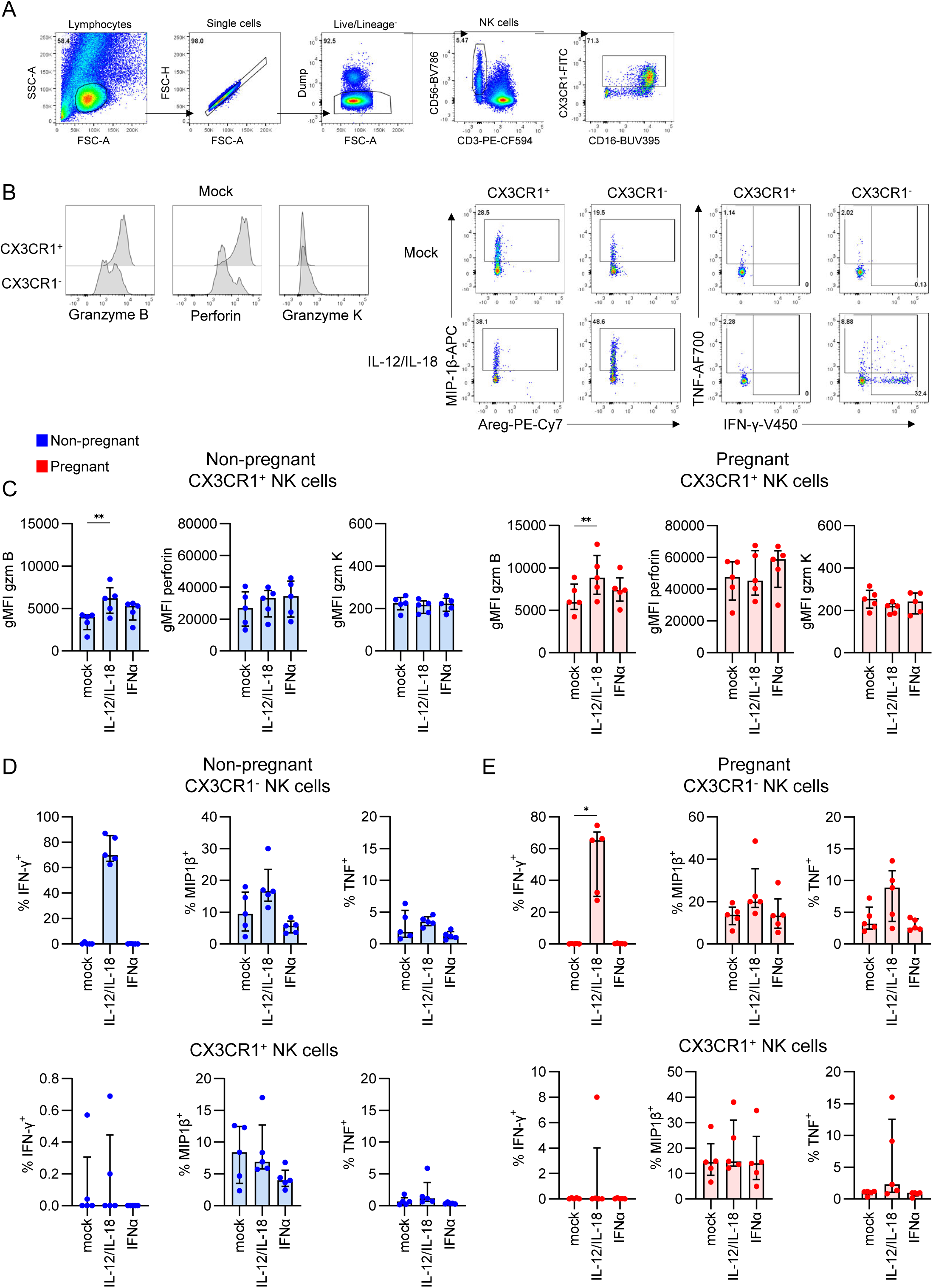
Activation profiles of CX3CR1^+^ and CX3CR1^-^ NK cells. (A) Gating strategy and (B) representative FACS plots of granzyme B, perforin, granzyme K, IFN-γ, MIP-1β and TNF on NK cells. (C-D) Frequencies of IFN-γ^+^, MIP-1β^+^ and TNF^+^ on CX3CR1^-^ and CX3CR1^+^ NK cells across mock, IL-12/IL-18, or IFNα stimulation in (C) non-pregnant and (D) pregnant blood. Statistics shown are a Friedman test with Dunn’s multiple comparisons test.

